# Quantifying Contraceptive Side-Effects: A Prospective Cohort Study of Symptom Burden, Risk Factors, and Daily Life Disruption in South-Central Ethiopia

**DOI:** 10.1101/2025.03.23.25324335

**Authors:** Rose Stevens, Eshetu Gurmu, Chris Smith, Rebecca Sear, Ametelber Negash, Virginia J. Vitzthum, Tamrat Abebe, Sisay Teklu, Jenny A. Cresswell, Elizabeth Ewart, Lemlem Kebede, Alexandra Alvergne

## Abstract

Unwanted side-effects are the leading cause of dissatisfaction and discontinuation of hormonal contraceptives worldwide. Yet contraceptive side-effects are commonly dismissed as minor and/or misconceptions within global health, in part due to the paucity of quantitative data on side-effects symptoms. This research aimed to (1) compare changes in symptom number and severity among hormonal contraceptive users and a control group over a 3-months period, (2) identify risk factors for such changes, and (3) evaluate their impact on women’s daily lives. We conducted an observational baseline-controlled prospective cohort study among injectable and implant users and a control group of non-users in Central Oromia, Ethiopia. Sociodemographic, diet, activity data and monthly side-effect symptoms were collected from pre-initiation to three months. Multilevel models adjusted for temporal autocorrelation were used to evaluate change in the number and severity of symptoms. Minimally adjusted models were used to identify risk factors for increased negative symptoms among contraceptive users and evaluate the impact of experiencing symptoms on women’s daily activities. A total of 278 participants (106 injectable, 72 implant, 100 non-users) were included for analysis. Compared to pre-initiation, injectable users experienced 28% more symptoms at month 3 (adjusted incident rate ratio (IRR): 1.28, 95% CI: 1.05 – 1.57 p = 0.015), implant users experienced a peak of 41% more symptoms at month 2 (adjusted IRR 1.41, 95% CI: 1.15 – 1.73, p = 0.002), and non-users experienced no changes over a similar time period. Contraceptive users with physically demanding occupations, food insecurity, and a history of recent infection experienced the greatest symptom severity, also associated with negative impacts on women’s activities, including work, chores, and relationships. These findings indicate that reducing the burden of contraceptive side effects requires addressing underlying health stressors and considering the significant impact of side-effects on women’s daily lives, rather than relying solely on dispelling misconceptions.

**Key messages:** *What is already know on this topic:*

- Existing research lacks the data necessary to both identify risk factors for contraceptive side effects and assess the extent of daily disruptions caused by these symptoms.

*What this study adds:*

- The design of this study enables us to demonstrate that side-effects are: (1) significant: users report an increase in symptoms after initiating contraception, unlike non-users who do not exhibit such changes; (2) predictable: women experiencing health stressors (nutritional, physical and infectious) prior to initiation report the greatest number and severity of side-effects when using hormonal contraception; (3) disruptive: higher symptom severity is associated with a decreased ability to carry out key daily activities pertaining to work, relationships, and house chores.

*How this study might affect research, practice and/or policy:*

- Contraceptive counselling should be sensitive to variation in risk of side-effects and support women with high symptom burdens with management options or method switching, rather than dismissing concerns as misconceptions.
- Our findings highlight the need for further research confirming predictive drivers of side-effect experiences to guide counselling and to move towards personalised contraceptive technology development.

## Introduction

Many women worldwide want to use hormonal contraception but often hesitate or forego its use due to anticipation or prior experience of side-effects (1,2). Qualitative studies have documented side-effects with significant impacts on women’s lives, including irregular bleeding, nausea, headaches, slow returns to fertility, mood changes, and loss of libido amongst others (3,4), yet the dominant global health discourse assumes that many side-effect concerns are myths, or at worst, relate to minor symptoms (5) - which can be alleviated through counselling alone (6,7). This framing of fears as exaggerated and dismissible is not uncommon for health issues which typically affect women, particularly if they affect groups of already marginalised women (8,9), and this gender bias extends into wider lack of research into women’s health and pain more generally (10,11). Additionally given the limited evidence supporting the effectiveness of counselling strategies aimed at dispelling so-called myths about contraceptive side-effects (12), there is a clear need for a quantitative assessment of side-effect experiences, coupled with a robust epidemiological design in settings representative of the environments in which women typically use hormonal contraception.

Currently, the most widely used and standardised data source on contraception in low- and middle-income countries (LMICs), the Demographic and Health Survey (DHS), measures the prevalence of ‘fear of side-effects or health concerns’ among past users of contraception. This category has been critiqued as too broad and non-specific (13,14), lumping all side-effect experiences and fears together, and failing to measure the occurrence of any side-effects experiences among current users. Since 2020, studies measuring side-effects symptom experiences have emerged within population health (15–19), but no standard-use validated measure for contraceptive side-effect symptoms is available. Those measures that do exist often capture a limited range of symptoms without considering symptom severity or impact on women’s lives (17,20) and are not designed to be used over time (but see 18,19). Further, existing studies lack a baseline measure of symptoms before use and a control group of non-users, making it difficult to attribute negative symptoms to hormonal contraceptive use. Given the limitations of current methods for measuring side-effects, several studies have called for “further methodological research to identify how best to accurately identify and quantify the experience of side-effects in large-scale, population-based surveys” (18).

The risk of experiencing side-effects and variation in the tolerability of hormonal contraceptives may be predictable (21–23). Reproductive ecologists Vitzthum and Ringheim (2005) posited that those living with nutritional, physical, and immune stress, who typically show reduced reproductive hormone levels (24), may be the least able to tolerate high doses of exogenous reproductive hormones. There is indirect support for this hypothesis from Ethiopia from both qualitative studies documenting the perception that women who experience food insecurity and heavy workloads are those who suffer most (3,25) and from an analysis of the Ethiopian Demographic and Health Survey showing increased risk of discontinuation of the injectable contraceptive due to side-effects among anaemic women (26). The hypothesis according to which women vary in their risk of experiencing contraceptive side-effects has yet to be tested using quantitative side-effect measures.

Ethiopia provides a relevant setting for understanding variation in contraceptive side-effects because contraception is widely available to those who desire to use it due to a well-established family planning program. Yet, the proportion of women who are categorized as having an unmet need for contraception remains high, in large part due to health concerns and fear of side-effects (27), and there remain large urban-rural inequalities (16% unmet need in rural areas; 9% in urban areas) (28). Most Ethiopian contraceptive users use one of two methods: the 3-month progestin injectable contraceptive, and increasingly the 3-year progestin implants, which make up 65% and 21% of modern contraceptive use respectively (29). Both methods have been associated with high side-effects burdens (30,31). This narrow method mix makes it easier to adjust for variability in side-effects introduced by method type.

The aims of this prospective cohort study in South Central Ethiopia were three-fold: (1) to test the hypothesis that the number and severity of side-effect symptoms increase after initiating hormonal contraception compared to both before initiation and a control group of non-users, (2) to evaluate if risk factors indicative of life conditions with high physical, nutritional, and immune stress showed an association with increased side-effects among users and (3) to quantify the impact of contraceptive side-effects on the ability of users to carry out different daily activities. The data are relevant for informing strategies to reduce the burden of hormonal contraceptive side-effects.

## Methods

This is a prospective observational cohort study among injectable and implant users and a control group of non-users in Central Oromia, Ethiopia. We selected 4 urban districts (kebeles) in the city of Adama in the East Shewa zone of Oromia and 5 surrounding rural districts in Adama county to capture a range of contexts. Two health extension workers (HEWs) were hired in each district to undertake recruitment and data collection. Data collection occurred between May and December 2021. HEWs explained the study to potential participants and obtained written informed consent from those wishing to participate. The period of follow-up was 3 months and women were compensated with a kilo of coffee as a gift in kind for participation. All data collectors were provided with masks and hand sanitiser for themselves and participants during data collection.

Ethical approval was obtained from three bodies: the Oxford Tropical Research Ethics Committee (OxTREC) at the University of Oxford (562-18); the College of Health Sciences at Addis Ababa University (069/19/DMIP), and the Oromia Health Bureau (BEFO/HBTF/1-16/239). Letters of permission were obtained from health offices at the zonal and woreda (county) levels and administrating health centre at the kebele level.

### PPI statement

Patients and public were involved in research design and method development. First, a stakeholder workshop with policy makers, NGO representatives, and health professionals was held in Addis Ababa prior to data collection to obtain feedback on research priorities and study methodology. Secondly, prior qualitative discussions with contraceptive users allowed us to incorporate the voices and priorities of women in the local area through exploring which questions to prioritise and informing the development of a context-specific side-effect measurement instrument. This design responds to calls from the field of evidence-based medicine to increase qualitative research into patients’ priorities and their experience of illness across different circumstances (56).

### Study population

Women aged 20-30 who were naturally cycling with semi-regular menstrual cycles (between 21 and 45 days), who had not used the injectable or implant in the past 6 months nor any other contraceptives/long term medication in the past 3 months and who were not pregnant or exclusively breastfeeding were included in the study. Those seeking to begin the injectable or implant were recruited into the contraceptive groups. Non-users fitting the inclusion criteria were recruited from the general population at or nearby the health facility. A realistic power calculation was not possible due to the limited published data on contraceptive side-effects variance and recruitment figures were calculated based on cost and time constraints.

### Instrument for measuring contraceptive side-effects

We developed a locally specific *Symptom Questionnaire (SQ)* based on qualitative interviews and focus group discussions with contraceptive users in the East Shewa zone (32). It captured self-reported presence and severity of 25 different symptoms over the last month, as well as the impact of these symptoms upon 7 different day-to-day activities (online supplemental file 1). The SQ was designed to be used prospectively, asking about symptoms in relation to the last month, reducing recall bias and capturing incident symptoms soon after they were experienced. The SQ focuses on ‘symptoms’ rather than ‘side-effects’ to allow its use with non-users and to prevent contraceptive users from being influenced to report more symptoms, although blinding to contraceptive use was not feasible.

### Procedure

The *Symptom Questionnaire (SQ)* was administered at recruitment and then monthly for each of the 3 following months to all participants. At recruitment, none of the participants were using hormonal contraception; initiation of either the implant or injection started within a week of recruitment for the user groups. During follow up, each *SQ* was collected for all participants as close as possible to a month after the last measure (number of days between measures: median (IQR): 30 (30;35)). Study staff collected paper *SQ* from HEWs once a week and this data was double entered in both Ethiopia and the UK; disparities were checked against original surveys. To collect data on risk factors, a *Socio-Demographic Survey (SDS)* was administered by a research assistant who visited each woman within a month of recruitment. The SDS included questions on sociodemographic profile, diet, activity, health, and reproductive history (online supplemental file 2).

### Statistical analysis

#### (1) Changes in symptoms number and severity over time in users and non-users

Our primary outcome was **the number of side-effect symptoms** reported in the last month. Our secondary outcome was **the summed total of all symptom severities (symptom severity score)** in the last month, with each symptom contributing a greater score to the total depending on how severely it was experienced (0: not experienced, 1: mildly, 2: moderately, 3: severely). To evaluate differences in symptom trajectories between contraceptive users and non-users, we ran a multilevel and multivariable analysis. We applied a Poisson distribution to account for the skewed distribution of count data. The multilevel structure of the model accounted for the clustered data, i.e. individuals recruited in the same district and by the same HEW. The variable month was included as a random slope to account for temporal autocorrelation. Month was also included as a fixed categorical variable due to the small number of time points considered. An interaction term between month and contraceptive group was included as a fixed effect to allow for the differential effect of time between non-users and users. We reported crude and adjusted incident rate ratios (IRR), capturing the change in incidence of symptoms over time, 95% confidence intervals and p values to determine the strength of the association between contraceptive use and side-effect symptom changes. A principle component analysis (PCA) was conducted to investigate which symptoms were reported together in users (after initiation) and non-users (33). We conducted sensitivity analyses to (i) evaluate the contribution of each symptom to overall trends and (ii) assess the robustness of the findings by excluding mild symptoms.

#### (2) Risk factors for symptom severity scores after three months of contraceptive use

For each risk factor , we ran a minimally adjusted Poisson model. We adjusted for the difference between the individual and the mean baseline scores to account for those reporting higher symptom severity scores at baseline and to minimise any effects of regression towards the mean (34). All continuous exposure variables were scaled and were tested as linear or quadratic terms. Akaike Information Criterion (AIC) was used to compare models fit. We accounted for multiple testing using the False Discovery Rate (Benjamni-Hochberg procedure). We undertook sensitivity analyses by (i) restricting the data to symptoms reported by users after two months, and (ii) repeating the analysis among non-users to assess the specificity of risk factors related to the group of contraceptive users. Given the non-randomised study design, we avoid drawing conclusions about risk factors for side-effects based on differences between contraceptive user and non-user groups and focus on providing exploratory evidence for side-effect risk factors among users.

#### (3) Associations between symptoms severity and women’s daily lives

We modelled whether women’s ability to undertake six different daily activities in the last month was negatively affected by her symptoms (yes/no), as reported by participants, as a function of the number and severity of contraceptive users’ symptoms, using logistic regression models adjusted for relevant confounders. For each activity, we ran a set of 25 minimally adjusted models to explore which symptoms were driving any observed associations. Because several symptoms commonly co-occur (online supplemental file 3), these associations are not independent.

#### (4) Causal inference

Adjustment variables were chosen based on directed acyclic graphs (online supplemental file 4) (35). Whilst causal inference is difficult in observational studies, scholars have argued that there is merit in accepting our intention to investigate causality, making our assumptions and limitations clear, and not shying away from the causal goal of research (36). All analysis was undertaken in R version 4.2.3.

## Results

A total of 368 women were recruited, of which 278 were considered in our analyses based on the availability of side-effect data at all time points and meeting the inclusion and exclusion criteria (Figure 1, online supplemental file 5). Only 0.3% of data in risk factors and covariates was missing, with a maximum of 3% missing in any one given variables, thus we analyzed complete cases. Socio-demographic and lifestyle characteristics were broadly comparable between those included and those excluded from the final sample (online supplemental file 6), although individuals excluded had higher socioeconomic scores, were older, had higher parity, and were more likely to be urban and not undertake vigorous work. Excluded individuals also had higher mean number of symptoms in months 2 and 3. This suggests that the results underestimate the association between contraceptive use and increased symptom scores, reducing risk of Type 1 error from their exclusion.

**Figure 1.**
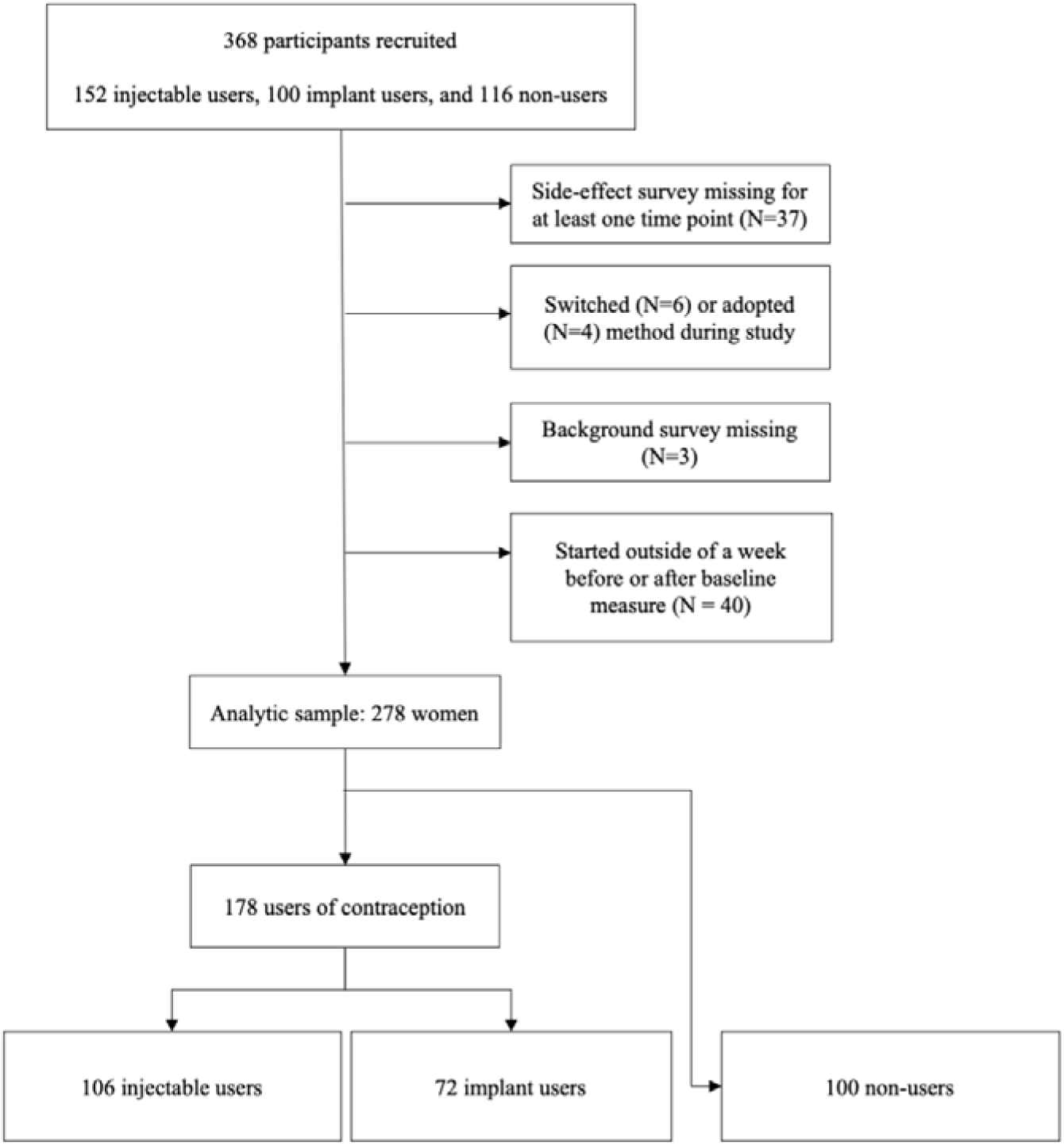
Analytic sample flowchart

In this sample, contraceptive users were older, more likely to be housewives and be married, less educated, more likely to be a Christian Orthodox and to have children (online supplemental file 7). There was minimal difference between contraceptive users and non-users in the number or symptom severity scores experienced at baseline (pre-initiation for contraceptive users). The mean number of symptoms experienced in the last month at baseline was 3.8 (SD 3.4) and 3.8 (SD 3.2) among injectable and implant users respectively, and 4.0 (SD 3.6) among non-users. Injectable users’ symptoms increased over time to peak at 4.2 (SD 2.9) after 3 months of use. Implant users’ symptoms increased to 4.5 (SD 3.3) symptoms after two months of use, and then decreased again by month three. While these mean symptom increases are modest, they mask much variation. Those in the top decile of the number of symptoms experienced after three months of contraceptive use had a mean increase of 6.2 (SD 4.1) symptoms from pre-initiation to month 3 (mean at baseline = 4.5 (SD 3.6) and mean at month 3 = 10.7 (SD 1.2)), among both the injectable and the implant. Among non-users, symptoms decreased over time, dropping to 2.6 (SD 2.3) by month three.

After 3 months of use, the most common symptoms among both injectable and implant users were not having a period, back and joint pain, fatigue, and headache (Figure 2). There was variation in which symptoms changed most over time. From baseline to month 3, injectable users showed the greatest increase in fatigue, not having a period, increased appetite, fever, and back and joint pain; implant users showed the greatest increase in not having a period, sleep disturbance, fatigue, reduced sex drive or enjoyment, and face marks (Figure 2, online supplemental file 8). Non-users on average reported reductions in symptoms over time with a particular reduction in proportions experiencing dizziness and fever, due to the high proportions (∼ a third) of non-users reporting a cough in the three months prior to recruitment, potentially indicating higher levels of illness upon recruitment and decreases in symptoms over recovery. The occurrence of fever at baseline may be driven by the recruitment strategy used by HEWs, who selected participants among visitors to health centres and health posts, which, for the non-user group, was likely composed of people attending due to existing illness.

**Figure 2.**
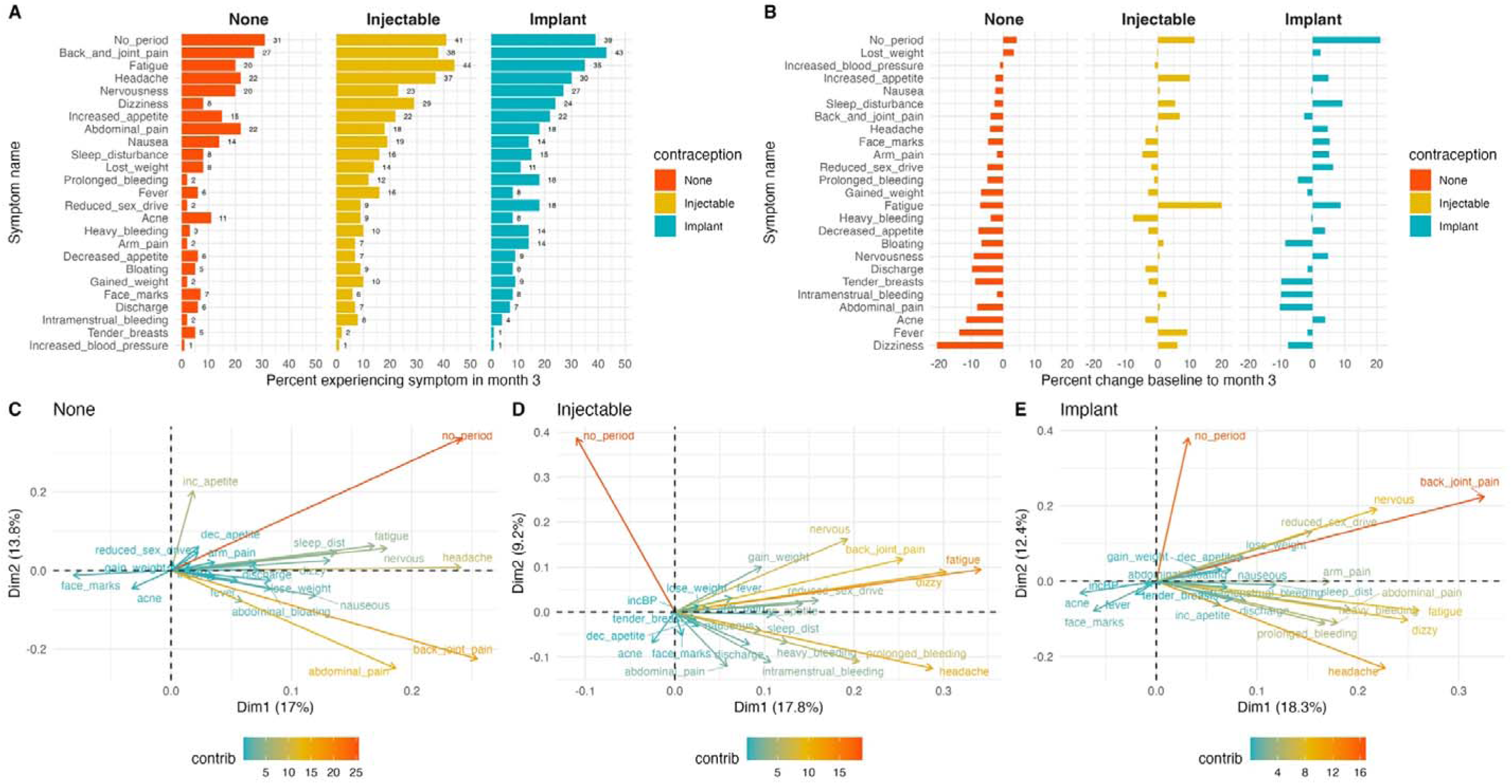
(A) Percent experiencing each symptom among users and non-users of contraception after 3 months (B) Percent change in proportion experiencing in each symptom among users and non-users of contraception from baseline to after 3 months (C, D, E) Biplots showing results of a principle component analysis of symptom co-occurrence in all months (excluding baseline) for non-users, injectable, and implant users respectively. Similar direction of arrows in C, D, and E shows positive correlation of symptoms and length of arrows shows the importance of that symptom in explaining variance in responses.

We found no clear distinct dimensions of symptoms among contraceptive users, with the exception of a second ‘no period’ dimension, independent from most other symptoms (Figure 2). Most symptoms covaried along the first dimension, suggesting that the presence of one symptom was associated with the presence of others, providing validity to our approach modelling total symptom counts. Fatigue, dizziness, headache, back and joint pain contributed most variation in the symptom set.

### (1) Changes in symptom number across groups

Non-users did not show a statistically significant change over time in the number of symptoms they experienced. By contrast, users of the injectable showed a greater incidence of symptoms in months 1, 2, and 3 compared to pre-initiating the injectable. Implant users similarly showed greater incidence of symptoms in months 1 and 2 compared to pre-initiation (Table 1). These associations are adjusted for age, socioeconomic status, parity, urban/rural residence, and fever at baseline. Injectable users’ symptom incidence was 28% higher at month 3 (adjusted IRR (aIRR) 1.28, 95% CI: 1.05 – 1.57, p=0.015), equating to approximately an extra two thirds of a symptom compared to pre-initiation. Implant users’ symptom incidence was 41% higher at month 2 (aIRR 1.41, 95% CI: 1.15 – 1.73, p=0.001), equating to approximately an extra one symptom compared to pre-initiation. This was followed by a slightly lower incidence in month 3 among implant users which did not differ significantly from pre-initiation (aIRR 1.21: 0.95 – 1.55, p=0.116).

**Table 1:** Crude and adjusted incident rate ratios for the impact of contraceptive use on symptom number and severity over time. Estimates adjusted for age, socioeconomic status, parity, rural/urban residence, and fever at baseline in interaction with time. Each column is derived from the same models using a different reference group. (a)IRR = (adjusted) incident rate ratio. CI = confidence intervals. Bold values represent p<0.05.

| \|Group: | None |  | Injectable |  | Implant |  | None |  | Injectable |  | Implant |  |
| --- | --- | --- | --- | --- | --- | --- | --- | --- | --- | --- | --- | --- |
|  | IRR (95% CI) | p value | IRR (95% CI) | p value | IRR (95% CI) | p value | aIRR (95% CI) | p value | aIRR (95% CI) | p value | aIRR (95% CI) | p value |
| Crude estimates |  |  |  |  |  |  | Adjusted estimates |  |  |  |  |  |
| Symptom number |  |  |  |  |  |  |  |  |  |  |  |  |
| Intercept | 2.93 |  | 3.07 |  | 3.36 |  | 1.93 |  | 2.22 |  | 2.40 |  |
| Month 1 | 0.97 (0.83 – 1.14) | 0.704 | 1.12 (0.96 – 1.30) | 0.137 | 1.20 (1.01 - 1.43) | 0.043 | 1.10 (0.92 – 1.32) | 0.294 | 1.18 (1.02 – 1.37) | 0.031 | 1.27 (1.06 – 1.51) | 0.009 |
| Month 2 | 0.77 (0.63 – 0.95) | 0.013 | 1.23 (1.03 – 1.47) | 0.022 | 1.28 (1.04 – 1.58) | 0.022 | 1.00 (0.81 -1.24) | >0.9 | 1.32 (1.11 – 1.56) | 0.002 | 1.41 (1.15 – 1.73) | 0.001 |
| Month 3 | 0.71 (0.56 – 0.91) | 0.006 | 1.18 (0.95 – 1.46) | 0.128 | 1.08 (0.84 – 1.41) | 0.541 | 0.92 (0.72 – 1.18) | 0.528 | 1.28 (1.05 – 1.57) | 0.015 | 1.21 (0.95 – 1.55) | 0.116 |
| Symptom severity score |  |  |  |  |  |  |  |  |  |  |  |  |
| Intercept | 3.54 |  | 3.56 |  | 4.10 |  | 2.46 |  | 2.89 |  | 3.30 |  |
| Month 1 | 0.89 (0.70 – 1.14) | 0.373 | 1.22 (0.97 – 1.54) | 0.085 | 1.29 (0.98 – 1.70) | 0.064 | 1.14 (0.96 – 1.35) | 0.128 | 1.29 (1.12 – 1.49) | 0.001 | 1.38 (1.17 – 1.63) | <0.001 |
| Month 2 | 0.73 (0.55 – 0.95) | 0.021 | 1.35 (1.05 – 1.72) | 0.018 | 1.30 (0.97 – 1.74) | 0.083 | 1.07 (0.85 -1.35) | 0.567 | 1.45 (1.20 – 1.76) | <0.001 | 1.46 (1.17 – 1.84) | 0.001 |
| Month 3 | 0.73 (0.54 – 0.98) | 0.039 | 1.37 (1.04 – 1.81) | 0.024 | 1.15 (0.83 – 1.61) | 0.393 | 1.00 (0.74 – 1.34) | >0.9 | 1.40 (1.09 – 1.80) | 0.008 | 1.19 (0.88 – 1.61) | 0.266 |

### (2) Changes in symptom severity across groups

We found stronger evidence for an increase in severity scores among contraceptive users over time (Figure 3, Table 1) than when predicting number of symptoms alone. In models adjusted for age, socioeconomic status, parity, rural/urban residence, and fever at baseline, the incidence of side-effects weighted by their severity did not change with time for the non-user group, but it was 45% higher for injectable users (aIRR 1.45, 95% CI: 1.20 – 1.76, p<0.001) and 46% higher for implant users (aIRR 1.46 ,95% CI: 1.17 – 1.84, p=0.001) after two months of use compared to pre-initiation. A sensitivity analysis excluding mild symptoms showed the same trends, as did removing the multilevel structure of the data. Using individual logistic regression models for the occurrence of each symptom over time, we found only a few symptoms with strong associations with contraceptive use (online supplemental file 9). Increased appetite and fatigue were associated with injectable use while not having a period, acne, discharge, headache, and fatigue were associated with implant use. These findings suggest either that our study is underpowered to detect symptom level trends or that contraceptive use is not consistently associated with a particular type of symptom. This latter explanation fits with the weak correlation coefficients between most symptoms (online supplemental file 3).

**Figure 3.**
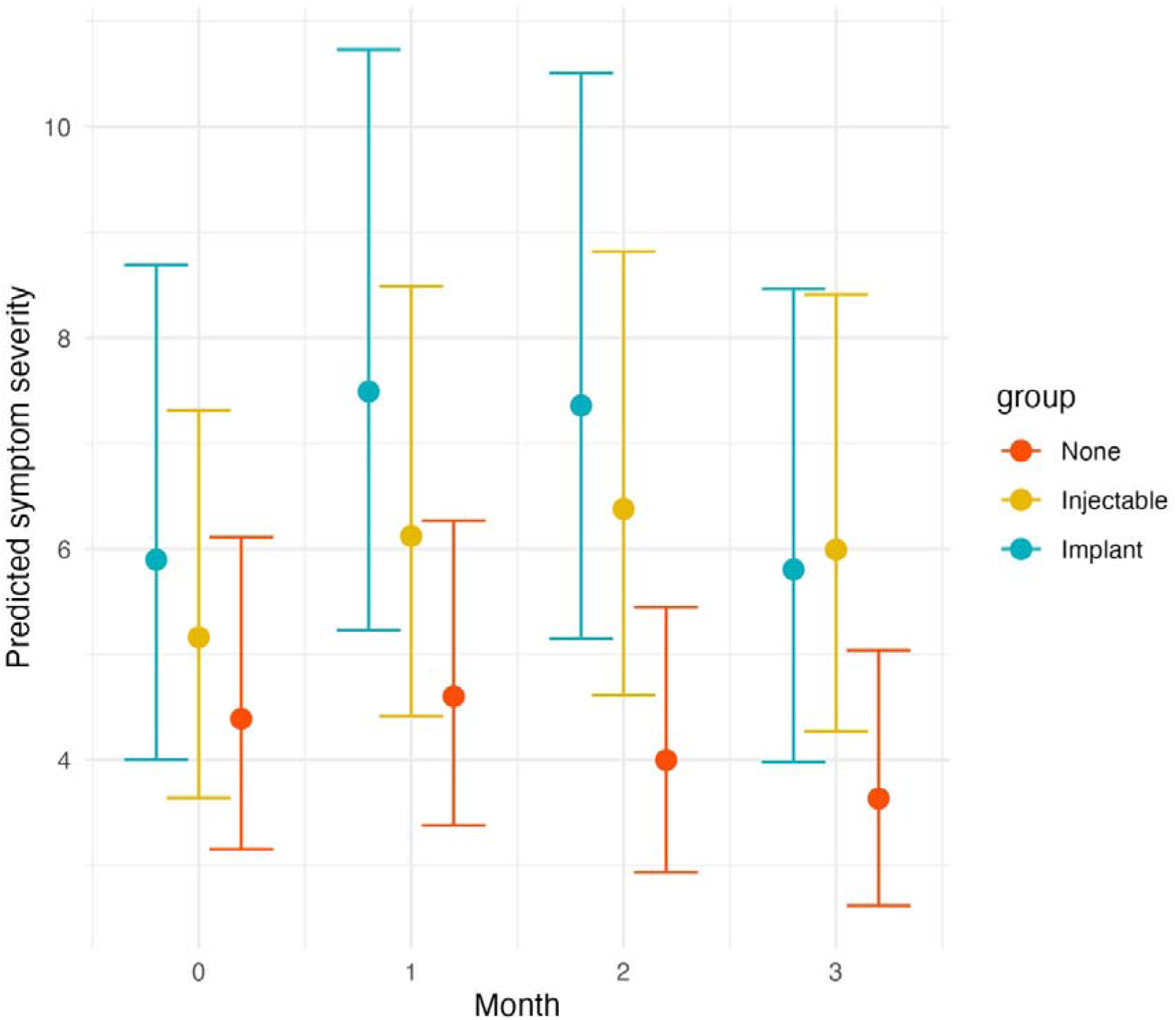
Predicted symptom severity scores over time across groups. Predicted summed severity scores and 95% CIs of symptoms experienced by each contraceptive group across time and adjusted for age, socioeconomic status, parity, rural/urban residence, and fever at baseline in interaction with time.

### (3) Risk factors for contraceptive side-effects

We tested independent associations between overall symptom severity score and 28 risk factors, accounting for multiple testing. Increased symptom severity score after three months of contraceptive use was independently associated with older age (IRR 1.11, 95% CI 1.05-1.18, FDR=0.002), urban residence (IRR 1.81, 95% CI 1.60-2.05, FDR<0.001), Muslim religion (IRR 1.88, 95% CI 1.60-2.20, FDR<0.001, ref = Christian Orthodox), higher/technical education (IRR 1.33, 95% CI 1.15-1.54, FDR<0.001, ref = no education), and being in the highest socioeconomic quintile (IRR 1.36, 95% CI 1.10-1.69, FDR=0.010, ref = lowest quintile) (online supplemental file 10). Adjusting for age, socioeconomic status, education, religion, and urban rural residence (online supplemental file 11), symptom severity scores increased with reproductive factors such as having ever been pregnant (aIRR 1.35, 95% CI 1.07-1.70, FDR=0.042), having 1 child (aIRR 1.52, 95% CI 1.22-1.91, FDR=0.001, ref = no children), or having had a child more than 6 months prior to the survey. Symptom severity scores increased with proxies of physiological stress including having an occupation that involves physical activity (aIRR 1.38, 95% CI 1.17-1.63, FDR=0.001, ref = housewife), having had symptoms of infection, such as a cough, diarrhoea, or fever in the 3 months prior to recruitment (aIRR 1.27, 95% CI 1.12-1.45, FDR=0.002), food security in the last four weeks: having had fewer meals than usual (IRR 1.34, 95% CI 1.14-1.58, FDR=0.002) or having gone to bed hungry (aIRR 1.47, 95% CI 1.14-1.91, FDR=0.015), and eating eggs less than once a week, representing poor dietary nutrition in terms of fat and protein (aIRR 1.32, 95% CI 1.10-1.57, FDR=0.011). Contrary to prediction, having eaten meat less than once a month was associated with lower symptom severity (aIRR 0.84, 95% CI 0.74-0.95, FDR=0.028). This result may be due to reverse causation or association between meat eating and infection. Meat consumption in is very low in this sample (15%), as across Ethiopia (37), limiting our ability to investigate the role of different frequencies of meat consumption. Finally body fat showed an inverse U-shaped relationship with symptom severity (body fat aIRR 1.62, 95% CI 1.22-2.17, FDR=0.006; body fat squared aIRR 0.56, 95% CI 0.42-0.76, FDR=0.001): participants with 20% body fat reported the highest severities, while participants with both higher (20% to 40%) and extremely low body fats reported lower symptom severity. Why extremely low levels of body fat associate with lower symptom severity is puzzling, and may result from measurement error or confounding (Figure 4, online supplemental file 11).

**Figure 4.**
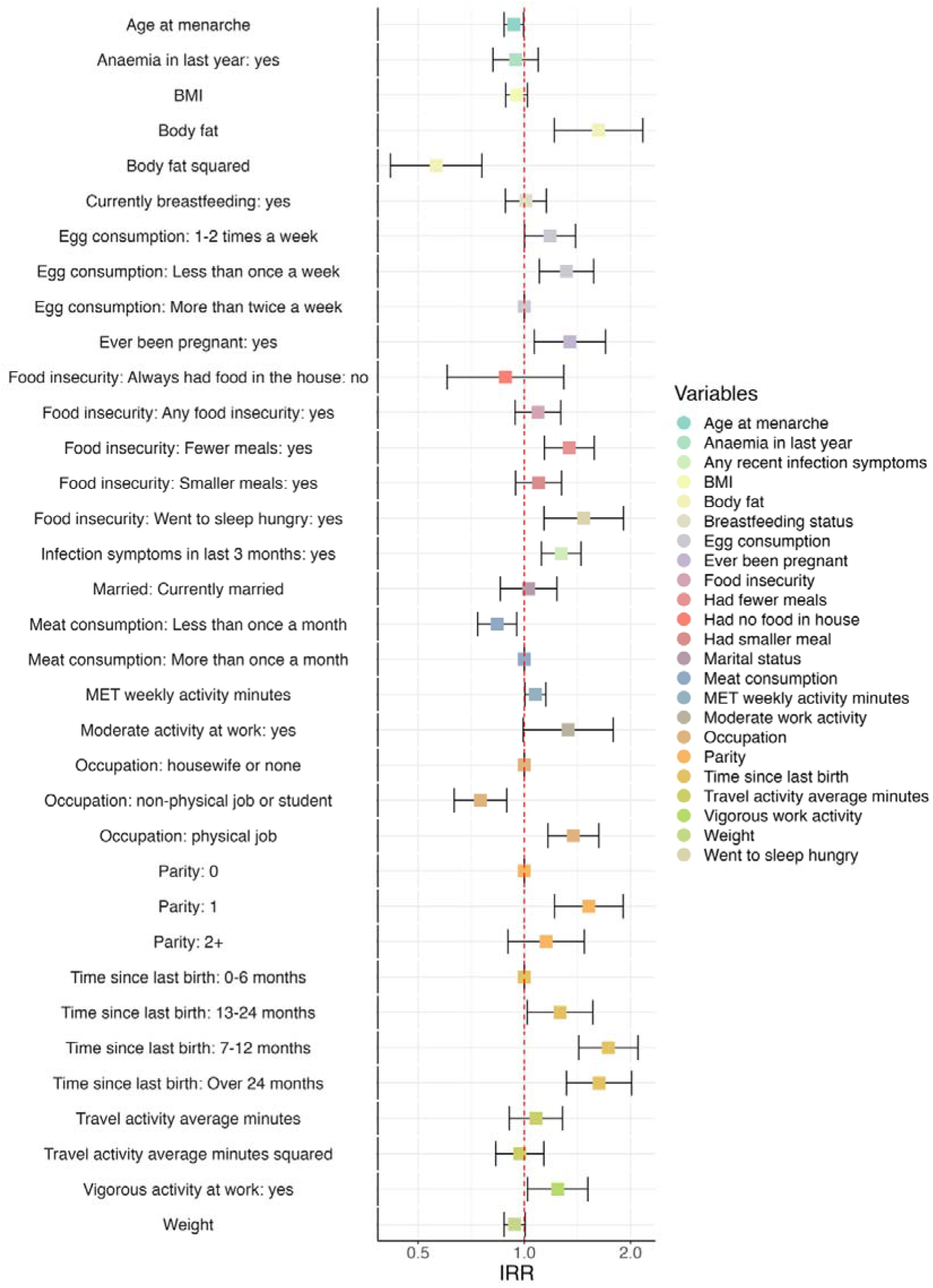
Forest plot showing adjusted association between risk factors and summed severity scores among users of progestin-based contraception at month 3. Results show incident rate ratios and 95% confidence intervals from minimally adjusted individual models for each risk factor using a Poisson distribution. Adjustment variables included age, urban/rural residence, education, religion, socioeconomic status, and the difference between individual baseline symptoms and mean baseline symptoms. References categories for non-binary variables are included for comparison. Body fat shows two estimates from the same model, as it was modelled quadratically.

Similar results were found in a sensitivity analysis investigating risk factors for symptom severity after two months of contraceptive use, when implant symptoms peaked (online supplemental file 12). Among stress factors, nutritional stress specifically was found to be specific to contraceptive users only and not non-users (online supplemental file 13).

### (4) Associations between symptom severity and ability conduct daily activities

We investigated the associations between symptom severity after three months of contraceptive use and the ability to conduct six daily activity types: employment, education, daily chores, childcare, having a harmonious relationship, and religious practice. Symptom severity score reported by implant and injectable users was associated with a lower ability to undertake all activities. This is at the exception of education, likely due to the small numbers of contraceptive users who undertook educational activities each month (online supplemental file 14). The largest effects were found for religious activities (adjusted OR (aOR) 1.31, 95% CI: 1.16 – 1.53, p<0.001), employment (aOR 1.23, 95% CI: 1.07 – 1.45, p=0.007), and childcare (aOR 1.19, 95% CI: 1.08-1.35, p=0.001).

To further understand how side effects might disrupt daily life, we analyzed associations between individual symptoms and daily activities (online supplemental file 15). The correlations suggest physiological, physical, and cultural pathways of disruption. Physiologically, symptoms indicative of hormonal metabolism disruption—such as amenorrhea, unintentional weight changes, reduced appetite, sleep disturbances, anxiety, breast tenderness, and decreased sexual drive and enjoyment—can affect energy levels and are associated with reduced capacity for performing daily chores, providing childcare, and maintaining relationship harmony. Physically, pain symptoms (in the arm and abdomen) negatively impact the ability to manage household chores, childcare, and religious practices. Culturally, heavy bleeding is linked to reduced participation in religious practices, while amenorrhea is associated with disruptions across all other daily activities.

## Discussion

In this analysis of 278 users and non-users of injectable and implant contraception in central Oromia, Ethiopia, we show that (1) hormonal contraceptive use is associated with an increase in the number and severity of negative bodily and psychological symptoms after 3 months of use compared to before use and a control group, (2) risk factors for severe hormonal contraceptive side-effects include having a physical occupation, food insecurity, poor nutrition, and recent symptoms of infection, and (3) higher symptom severity is associated with a decreased ability to carry out key daily activities pertaining to work, relationships and house chores. These results are in line with findings from qualitative studies which document perceptions that some groups of women are better able to tolerate doses of hormonal contraception than others (3,32,38,39). The findings challenge the prevailing paradigm according to which contraceptive side-effects are minimal, exaggerated, or even mistaken in their attribution to contraceptive use.

Contraceptive users report a greater number and severity of side-effect symptoms over three months, whilst there is no significant change in symptoms among non-users. Our study, which followed groups of contraceptive users and non-users over time and measured symptoms before contraceptive initiation, suggests that hormonal contraceptives likely cause more severe symptoms. However, because we didn’t use a placebo in the non-user group, we can’t completely rule out other factors and thus can’t make absolute claims about causation. The pattern of how symptom severity increases over time vary by method: injectable users show a stable increase in symptoms over the three months period while implant users show a peak in symptoms after two months with a drop off by three months, supporting clinical guidance that some side-effects, particularly bleeding side-effects, might settle down after three months (40,41). Whilst the mean increase in number of symptoms observed was modest in absolute terms, this masked much variation and, for some, the increase was much greater. These findings highlight the blind spots of studies focusing entirely on mean increases of individual symptoms among all groups of women for drawing conclusions on the risk or seriousness of side-effects (23).

Counter to prediction, urban, richer, and more educated women report more severe side-effects in this sample. It is possible that this group of women report more severe symptoms due to greater awareness of health issues and lower normalisation of general poor health. For instance, urban educated women in Ghana were found to be particularly concerned about side-effects arising from hormonal contraception, as it jeopardised their ability to simultaneously achieve employment goals and become a mother when desired (42). Negative views of ‘modern’ contraception by educated women have been conceptualised as postmodern choices defying medicalisation (43) and health ideologies classing pharmaceuticals as harmful can emerge as a form of resistance to established hegemony (44). Urban educated women may therefore have a bias towards reporting more side-effects as part of a rejection of western contraceptives and increased concern about the impacts on their health - ideas which rural women may be less exposed to.

Our results indicate that women with more physically demanding occupations, with higher levels of nutritional stress and recent illness experience report the greatest symptom severities after three months of contraceptive use. These relationships may be mediated by reduced endogenous hormone levels (21), as suggested by studies showing an association between vigorous exercise, nutritional insecurity, and increased inflammation with suppressed progesterone and estrogen levels (45–47). In our sample, frequently eating eggs is a protective factor against increased contraceptive side-effects, which may arise because eggs are one of the highest dietary sources of oestrogen, potentially mitigating some of the effects of hypoestrogenism induced by progestin-based contraceptive use (48).

The findings suggest that contraceptive side-effects take a toll on a woman’s ability to undertake daily activities, as shown by previous qualitative research (49,50). Pain side-effects showed the strongest associations with women’s daily lives. Experiencing arm pain was negatively associated with the ability to provide childcare, potentially through limiting a woman’s ability to carry her child. Additionally, heavy bleeding was associated with self-reported negative consequences for religious practice, supporting previous research showing bleeding can prevent particularly Muslim women from praying or participating in religious activities (51,52). Reduced sex drive and enjoyment, along with gaining weight, feeling nervous, and sleep disturbances, were negatively associated with a woman’s ability to live a harmonious spousal relationship, calling into question the utility of contraceptive use if the motivation for and enjoyment of sex is reduced or if contraception negatively impacts a romantic relationship. The findings highlight the importance of including discussions around sexual satisfaction in family planning policy (53,54).

## Strengths and limitations

This is the first community-based observational study to prospectively collect data on contraceptive side-effect symptoms before and after initiation and among a concurrent group of non-users (19,55). It also utilised a locally-specific quantitative instrument developed specifically for this study. However, assigning any increase in negative symptoms to hormonal contraception is not fully possible because (i) the groups are not directly comparable due to non-randomisation and non-blinding participants to contraceptive use, and (ii) potential ceiling effects reduce the ability of our instrument to detect symptom changes among those with already large numbers of severe symptoms. Additionally, reporting biases may drive increased symptom levels among contraceptive users if they anticipate a greater number of symptoms, although this bias is unlikely to fully explain association with specific risk factors. Our results are limited in their generalisability because our side-effect instrument is specific to the population, but the results may be relevant to contraceptive users across other sub-Saharan African socioecological contexts with similar contraceptive method mixes and where levels of agricultural labour and food insecurity are high.

### Implications for practice

Contraceptive side-effects are an equity issue. Current recommended counselling is uniform across groups of potential users and which side-effects are dismissed as minor is determined with one broad brush approach, typically based on Western biomedical values and lifestyles (57). Our results bring into question the ethics of this approach as we find that women vary in which symptoms they experience, how much symptom severity changes over time whilst using contraception and how much symptoms jeopardise women’s ability to carry out their daily activities. Our findings suggest that women who already experience high burdens of physical, nutritional, and immune stress are those likely to report the most side-effects and the greatest impacts on their day-to-day activities.

## Conclusion

Contraceptive users experienced an increase in the number and severity of negative symptoms experienced across the first three months of contraceptive, while non-users did not. Risk factors for increased symptom severity over time with hormonal contraceptive use include physical stress, food insecurity, and recent poor health. Greater symptom severity was associated with negative impacts across many dimensions of women’s lives. While current recommended counselling is uniform across groups of potential users and side-effects are dismissed as minor (57), this study shows that contraceptive side-effects are both predictable and major.

## Additional information

### Contributions

Rose Stevens, Alexandra Alvergne and Eshetu Gurmu developed a proposal for the study. Rose Stevens, Alexandra Alvergne, Eshetu Gurmu, Chris Smith, Rebecca Sear, Tamrat Abebe, Sisay Teklu, Jenny Creswell, Virginia Vitzthum, and Elizabeth Ewart provided input for the study design. Rose Stevens, Alexandra Alvergne, Eshetu Gurmu, Chris Smith, and Ametelber Negash attended a stakeholder meeting in Addis Ababa with academics and policy makers to refine the methodology of the study. Ametelber Negash undertook the majority of the background data collection and supervised side-effects data collection by the health extension workers, with support from Rose Stevens and Eshetu Gurmu. Ametelber Negash’s role was taken over by Lemlem Kebede by the end of data collection. Eshetu Gurmu organised the data collection and data entry in Ethiopia. Rose Stevens undertook data analysis and manuscript writing with support from Alexandra Alvergne. Chris Smith, Virginia Vitzthum, Rebecca Sear and Ametelber Negash provided additional analysis review and support. This manuscript draft has been reviewed by all co-authors.

## Acknowledgements

We would like to thank all the Health Extension Workers in Adama City and Adama Woreda for their help following up participants in this study, and all the women who participated.

## Data Availability

Anonymised data underlying these analyses will be made available upon reasonable request.

