## Supplementary material for "Quantifying Contraceptive Side-Effects: A Prospective Cohort Study of Symptom Burden, Risk Factors, and Daily Life Disruption in South-Central Ethiopia": online supplemental file 1

### S1 Appendix: Side-effect measurement instrument

**Side-Effect Symptoms Tool**

Age ___ ___	Kebele _________________________	Date __________________________

Type of contraception using: None ____ Injectable _____	Implant _____ Date started contraceptive episode: ______________

Symptoms Module – Read out

We will now ask you some questions about different symptoms you may have experienced this month. Firstly, I will ask you how often you have experienced this symptom and then how severe was your experience. Finally, I will ask you how much this experience bothered you – was it a big problem for you or was it only minor bother?

| **(a) During the last month, did you experience…** | | | **(b)  How severe was it?** | | | **(c)   How much did this experience bother you?** | | | |
| --- | --- | --- | --- | --- | --- | --- | --- | --- | --- |
|  |  |  | **Mild** | **Moderate** | **Severe** | **Not at all** | **Little** | **Some-what** | **A lot** |
| Q1 increased appetite | O Most of the time | Answer parts (b) and (c) → | O | O | O | O | O | O | O |
|  | O Some of the time |  |  |  |  |  |  |  |  |
|  | O None of the time | ↓ Go to question 2 |  | | | | | | |
| Q2 decreased appetite | O Most of the time | Answer parts (b) and (c) → | O | O | O | O | O | O | O |
|  | O Some of the time |  |  |  |  |  |  |  |  |
|  | O None of the time | ↓ Go to question 3 |  | | | | | | |
| Q3 fever | O Most of the time | Answer parts (b) and (c) → | O | O | O | O | O | O | O |
|  | O Some of the time |  |  |  |  |  |  |  |  |
|  | O None of the time | ↓ Go to question 4 |  | | | | | | |
| Q4 raised blood pressure | O Most of the time | Answer parts (b) and (c) → | O | O | O | O | O | O | O |
|  | O Some of the time |  |  |  |  |  |  |  |  |
|  | O None of the time | ↓ Go to question 5 |  | | | | | | |
| Q5 fatigue | O Most of the time | Answer parts (b) and (c) → | O | O | O | O | O | O | O |
|  | O Some of the time |  |  |  |  |  |  |  |  |
|  | O None of the time | ↓ Go to question 6 |  | | | | | | |
| Q6 feeling dizzy or light headed | O Most of the time | Answer parts (b) and (c) → | O | O | O | O | O | O | O |
|  | O Some of the time |  |  |  |  |  |  |  |  |
|  | O None of the time | ↓ Go to question 7 |  | | | | | | |
| Q7 sleep disturbance | O Most of the time | Answer parts (b) and (c) → | O | O | O | O | O | O | O |
|  | O Some of the time |  |  |  |  |  |  |  |  |
|  | O None of the time | ↓ Go to question 8 |  | | | | | | |
| Q8 back or joint pain | O Most of the time | Answer parts (b) and (c) → | O | O | O | O | O | O | O |
|  | O Some of the time |  |  |  |  |  |  |  |  |
|  | O None of the time | ↓ Go to question 9 |  | | | | | | |
| Q9 pain in your arm | O Most of the time | Answer parts (b) and (c) → | O | O | O | O | O | O | O |
|  | O Some of the time |  |  |  |  |  |  |  |  |
|  | O None of the time | ↓ Go to question 10 |  | | | | | | |
| Q10 development of marks on your face (melasma/skin discolouration) | O Yes | Answer parts (b) and (c) → | O | O | O | O | O | O | O |
|  | O No | ↓ Go to question 11 |  | | | | | | |
| Q11 gaining weight | O Yes | Answer parts (b) and (c) → | O | O | O | O | O | O | O |
|  | O No | ↓ Go to question 12 |  | | | | | | |
| Q12 losing weight | O Yes | Answer parts (b) and (c) → | O | O | O | O | O | O | O |
|  | O No | ↓ Go to question 13 |  | | | | | | |
| Q13 not seeing your period (a missed period) | O Yes | Answer part (c) → |  |  |  | O | O | O | O |
|  | O No | ↓ Go to question 14 |  | | | | | | |
| Q14 feeling nervous or irritable | O Most of the time | Answer parts (b) and (c) → | O | O | O | O | O | O | O |
|  | O Some of the time |  |  |  |  |  |  |  |  |
|  | O None of the time | ↓ Go to question 15 |  | | | | | | |
| Q15 feeling nauseated | O Most of the time | Answer parts (b) and (c) → | O | O | O | O | O | O | O |
|  | O Some of the time |  |  |  |  |  |  |  |  |
|  | O None of the time | ↓ Go to question 16 |  | | | | | | |
| Q16 headaches or migraines | O Most of the time | Answer parts (b) and (c) → | O | O | O | O | O | O | O |
|  | O Some of the time |  |  |  |  |  |  |  |  |
|  | O None of the time | ↓ Go to question 17 |  | | | | | | |
| Q17 acne | O Most of the time | Answer parts (b) and (c) → | O | O | O | O | O | O | O |
|  | O Some of the time |  |  |  |  |  |  |  |  |
|  | O None of the time | ↓ Go to question 18 |  | | | | | | |
| Q18 swollen, tender or painful breasts | O Most of the time | Answer part (c) → | O | O | O | O | O | O | O |
|  | O Some of the time |  |  |  |  |  |  |  |  |
|  | O None of the time | ↓ Go to question 19 |  | | | | | | |
| Q19 lower abdominal pain (below the belly button) | O Most of the time | Answer parts (b) and (c) → | O | O | O | O | O | O | O |
|  | O Some of the time |  |  |  |  |  |  |  |  |
|  | O None of the time | ↓ Go to question 20 |  | | | | | | |
| Q20 lower abdominal bloating (below the belly button) (abdominal distension) | O Most of the time | Answer parts (b) and (c) → | O | O | O | O | O | O | O |
|  | O Some of the time |  |  |  |  |  |  |  |  |
|  | O None of the time | ↓ Go to question 21 |  | | | | | | |
| Q21 vaginal discharge (excluding bleeding) | O Most of the time | Answer parts (b) and (c) → | O | O | O | O | O | O | O |
|  | O Some of the time |  |  |  |  |  |  |  |  |
|  | O None of the time | ↓ Go to question 22 |  | | | | | | |
| Q22 heavy bleeding | O Most of the time | Answer parts (b) and (c) → | O | O | O | O | O | O | O |
|  | O Some of the time |  |  |  |  |  |  |  |  |
|  | O None of the time | ↓ Go to question 23 |  | | | | | | |
| Q23 prolonged bleeding (longer than my normal menses) | O Most of the time | Answer parts (b) and (c) → | O | O | O | O | O | O | O |
|  | O Some of the time |  |  |  |  |  |  |  |  |
|  | O None of the time | ↓ Go to question 24 |  | | | | | | |
| Q24 bleeding or spotting that was not my period (intramenstrual bleeding) | O Most of the time | Answer parts (b) and (c) → | O | O | O | O | O | O | O |
|  | O Some of the time |  |  |  |  |  |  |  |  |
|  | O None of the time | ↓ Go to question 25 |  | | | | | | |
| Q25 reduced sexual enjoyment and/or desire | O Most of the time | Answer parts (b) and (c) → | O | O | O | O | O | O | O |
|  | O Some of the time |  |  |  |  |  |  |  |  |
|  | O None of the time | ↓ Go to question 26 |  | | | | | | |

Now I will ask you some questions about your day to day activities to understand whether your symptoms affected your ability to carry out your activities. Firstly, I will ask about whether your carried out the activities, if you couldn’t do them because of your symptoms, or if you don’t do these activities normally. Then, if you did do it or couldn’t because of your symptoms, I will ask you whether about how much difficulty you had carrying them out due to the symptoms we discussed. Finally, I will ask you about how much any difficulty you had bothered you.

| **(a) During the last month, have you…** | | | **(b)  Due to the symptoms we have discussed, have you been able to carry out these activities…** | | | **(c)  Did any difficulty in carrying out this activity bother you …** | | | |
| --- | --- | --- | --- | --- | --- | --- | --- | --- | --- |
|  |  |  | **As usual** | **With some difficulty** | **With much difficulty** | **Not at all** | **Little** | **Some-what** | **A lot** |
| Q26 carried out your job (money generating activities/employment) | O Yes I carried out my job | Answer parts (b) and (c) → | O | O | O | O | O | O | O |
|  | O No I couldn’t carry out my job because of my symptoms |  |  |  |  |  |  |  |  |
|  | O No I didn’t do a job | ↓ Go to question 27 |  | | | | | | |
| Q27 undertaken some education (studying for a degree/qualification) | O Yes I attended my education | Answer parts (b) and (c) → | O | O | O | O | O | O | O |
|  | O No I couldn’t attend education because of my symptoms |  |  |  |  |  |  |  |  |
|  | O No I didn’t attend education | ↓ Go to question 28 |  | | | | | | |
| Q28 conducted daily chores (e.g. carrying items, cleaning clothes, grinding etc.) | O Yes I conducted chores | Answer parts (b) and (c) → | O | O | O | O | O | O | O |
|  | O No I couldn’t conduct any chores because of my symptoms |  |  |  |  |  |  |  |  |
|  | O No I didn’t conduct any chores | ↓ Go to question 29 |  | | | | | | |
| Q29 cared for your child/children | O Yes I cared for my children | Answer parts (b) and (c) → | O | O | O | O | O | O | O |
|  | O No I couldn’t care for my children because of my symptoms |  |  |  |  |  |  |  |  |
|  | O No I didn’t care for my children | ↓ Go to question 30 |  | | | | | | |
|  | O I don’t have children |  |  |  |  |  |  |  |  |
| Q30 had a harmonious relationship with my husband | O I am married and had a harmonious relationship | Answer parts (b) and (c) → | O | O | O | O | O | O | O |
|  | O I couldn’t have a harmonious relationship with my husband because of my symptoms |  |  |  |  |  |  |  |  |
|  | O I am married and could not have a harmonious relationship for other reasons | ↓ Go to question 31 |  | | | | | | |
|  | O I am not married |  |  |  |  |  |  |  |  |
| Q31 carried out religious practices (praying/attending a place of worship) | O Yes I carried out religious practice | Answer parts (b) and (c) → | O | O | O | O | O | O | O |
|  | O No I couldn’t carry out religious practice because of my symptoms |  |  |  |  |  |  |  |  |
|  | O No I didn’t carry out religious practice | ↓ Go to question 32 |  | | | | | | |
| Q32 been planning when and how many children to have | O Yes I have been planning when to have children | Answer parts (b) and (c) → | O | O | O | O | O | O | O |
|  | O No I have been unable to plan when to have children because of my symptoms |  |  |  |  |  |  |  |  |
|  | O No I have not been planning when to have children | ↓ End questions |  | | | | | | |
|  | O No I have not been planning when to have children |  |  |  |  |  |  |  |  |
