## Supplementary material for "Quantifying Contraceptive Side-Effects: A Prospective Cohort Study of Symptom Burden, Risk Factors, and Daily Life Disruption in South-Central Ethiopia": online supplemental file 2

### S2 Appendix: Variables included for analysis.

| **Variable** | **Measured** (questions, groupings, and analysis treatment) | **Justification** |
| --- | --- | --- |
| **Outcomes: Side-effects symptoms and impacts** | | |
| Symptom frequency | Symptom level– 1/0 from most, some, none of the time in the last month.  Survey level – summed number of symptoms experienced – “Symptom number” | Measure of whether a symptom was experienced or not. Covered 25 symptoms derived from qualitative discussions of reported side-effects among women in the local area. Summed number of symptoms allows a total score out of 25 for each survey as to the symptom burden experienced. |
| Symptom severity | Symptom level– 0-3 from not experienced, experienced mildly, experienced moderately, experienced severely. (Binary sensitivity analysis using 1/0 for moderately & severely vs mildly & not experienced.)  Survey level – summed number of symptoms experienced – “Symptom severity score” | Measure of how severely a symptom was experienced as qualitative discussion indicated wide variation in the severity or intensity of symptoms. Summed severity scores of symptoms allow a total score out of 75 for each survey as to the symptom burden experienced weighted by the severity of each symptom. |
| Impact on daily activities | Survey level - 6 activities. Binary variable 0/1 created from undertook the activity with no impact from symptoms (0) vs undertook the activity or could not undertake it because of the experienced symptoms. Those who would not otherwise undertake the activity anyway were coded as missing. | Measure of whether the overall symptoms level experienced in the last month had impacted 7 different daily activities, chosen based on qualitative discussions of side-effect impacts. |
| **Exposures** | | |
| **Contraceptive use** | | |
| Contraceptive use | Injectable user, implant user, or non-user. Participants recruited into one of 3 groups. Switchers and adopters excluded from analysis. | Injectable and implants are the most commonly used methods in Ethiopia, making up nearly 90% of modern contraceptive prevalence. Non-users were recruited as a control group. |
| **Socio-demographics** | | |
| Age | Continuous, in years | Hormone levels reduce nearer to menarche and menopause. Our inclusion criteria of 20-30 reduce risk of recruiting women near to these events. Age may also influence side-effect reporting. |
| Ecology | Rural; Urban. Measured at the level of kebele. Rural kebeles will be selected through characteristics such as predominant housing type, higher levels of food insecurity, high levels of agricultural labour and below average income levels of 4500 ETB per month or 54,000 ETB annually. Urban middle-class kebeles were selected based on high levels of sedentary/service-based occupations, access to main roads/urban centres, no food insecurity, relative affluence, high quality housing, and above average income levels of 4500 ETB per month or 54,000 ETB annually. | Measure of area of residence is predictive of many other sociodemographic, nutritional, and physical factors. Also, cultural variation in lifestyle and outlook may predict likelihood to report side-effects. |
| Education | None/Primary School; Secondary School; Higher education/technical training. Self-reported highest level of education attended. | Education is associated with choice of contraception and likely reported of side-effect symptoms. |
| Religion | Self-reported and categorised into Christian Orthodox, Muslim, Protestant and Other. Other include Waqefeta, a local traditional religion, and Catholicism. | Likely associated with contraceptive choice and possibly symptoms reporting. Associated with impact of symptoms, particularly bleeding symptoms. |
| Marital status | Self-reported as ‘married’ which comprised of legal marriage, traditional or informal, and cohabiting living as married. | Likely associated with contraceptive choice and possibly symptoms reporting. Associated with impact of symptoms. |
| Socioeconomic quintile | Quintiles of socioeconomic score (1 = low, 5 = high). Based on several variables (such as roof type or bank account ownership) transposed into a composite score using the ‘equity tool’, allowing calculation of a country specific measurement that places individuals into wealth quintiles (<http://www.equitytool.org/ethiopia/>). | Lower socioeconomic status has shown to be associated with lower progesterone levels in an urban Bolivian population (62) and may drive many of the other risk factors for side-effects. |
| **Physical stress** | | |
| Occupation type | Housewife or none; physical occupation (farmer, skilled manual labourer, unskilled labourer); non-physical occupation or student (professional, studying, trader). Based on self-reported categories. | Increased energy expenditure and physical activity has been associated with suppressed reproductive investment even in cases where there is not a calorie deficit (63). Participants in qualitative studies also report that those with hard physical workloads in their occupations are those that suffer most with side-effects. |
| Vigorous work | Self-reported that their work (including paid and unpaid work, household chores) contained vigorous activity (activities that require hard physical effort and cause large increases in breathing or heart rate, such as cutting crops, digging, shovelling, or grinding for at least 10 minutes continuously) | As above. All 4 metrics are derived from the World Health Organisation (WHO) standardised activity metric, the GPAQ tool (Global Physical Activity Questionnaire) (64). This asks about vigorous and moderate work and leisure activities and travel time, asking how many days a week each activity is undertaken and roughly how many minutes per day is spent in each activity. These activities are then weighted by their intensity and used to calculate. |
| Moderate work | Self-reported that their work (including paid and unpaid work, household chores) contained moderate activity (activities that require moderate physical activity and cause small increases in breathing or heart rate such as walking carrying light loads, planting and harvesting, washing, or tending to animals for at least 10 minutes continuously) |  |
| Travel activity time | A self-reported metric of how many minutes per week an individual spends walking or cycling in travel from place to place. |  |
| MET minutes | A metric of minutes of physical activity per week, weighted by the intensity of that activity. |  |
| **Nutritional stress** | | |
| Food insecurity | Binary summary of whether a participant answered yes to any of the following questions about the past four week: 1) have your family members eaten a smaller meal than you felt needed? 2) have your family members eaten fewer meals in a day than you felt needed? 3) have your family members had no food of any kind to eat in your household at least once? and 4) have your family members gone to sleep at night hungry? Each questions also included as a binary variable to investigate contribution of each. | Food security questions are based on a reduced set of the Household Food Insecurity Access Scale (HFIAS) (65). This gives a general indicator of nutritional and economic stress. |
| Frequency of egg and meat consumption | Participants were asked how often they ate 12 food groups in the last month. Response options were: more than once per day, once per day, 3–6 times per week, 1–2 times per week, 1–2 times per month and less than once per month. This measure was based on a similar food frequency questionnaire asked in a diet study in Hawassa, a city in southern Ethiopia. It was chosen for its concision (compared to diary measures or 24 hour recall), local applicability, and coverage of multiple food types and capture of frequency (66).  For eggs, these were grouped into more than once a week, 1-2 times a week, and less than once a month to create three roughly evenly sized categories. For meat, these were grouped into more than once a month, and less than once a month to create two roughly evenly sized categories. | Diet composition, particularly protein and fat levels, have been associated with hormone levels (67,68). Additionally, participants in qualitative studies report that side-effects may be mitigated by eating a high-quality diet, specifically referencing eating eggs and meat. |
| Anaemia | Self-reported measure of whether they had had anaemia in the last year. | Anaemic women previously have been shown to be at greater risk of discontinuation due to side-effects (26) and anaemia may indicate nutritional deficiencies or presence of infection (69) |
| **Health and Reproduction** | | |
| Recent symptoms of infection prior to use | Participants were asked if they had experienced a fever, a cough, or diarrhoea in the last three months. | Recent illness is predicted to be associated with suppressed progesterone levels and increased inflammation has been shown to be associated with lowered reproductive hormone levels (50). |
| Age at menarche | Self-reported in years and treated continuously | Earlier age of menarche is predicted to be associated with higher progesterone levels as earlier age of menarche has been associated with greater rates of ovulatory cycles and levels of ovarian steroid hormones (70). |
| Ever been pregnant | Self-reported. | May indicate general fecundity. |
| Parity | Measured through successive birth history. Categories into 0, 1, and more than 2. | Nulliparous women have been found to have lower progesterone levels (71). |
| Time since last birth | Self-reported months since last birth. Categorised into 0-6 months, 7-12 months, 13-24 months ago, over 24 months ago. | Recent birth may be associated with lowered reproductive hormone levels after birth and increased depletion of maternal resources in general, and thus increased susceptibility to side-effects. |
| Breastfeeding status | Self-reported as to whether participant is currently breastfeeding. Frequency of breastfeeding was also collected but not included in this variable. | As breastfeeding women do not return to cycling as quickly as non-breastfeeding women, due to the suppression of GnRH pulses among breastfeeding women. Thus, they maintain low levels of progesterone postpartum (72), it is predicted that breastfeeding women will have lower progesterone levels. Additionally, breastfeeding is very energetically costly and so may be associated with suppressed reproductive investment. |
| **Anthropometrics** | | |
| Weight | Measured using a Tanita BC-730 body composition monitor using single frequency bioelectrical impedance (BIA) scale and recorded in kilograms. Used with height (measured in centimetres) to calculate body mass index (BMI). | Weight has been associated with higher progesterone levels, but this relationship is not clear and may be inverse U-shaped (62,73). |
| Body fat percentage. | Measured using a Tanita BC-730 body composition monitor using single frequency bioelectrical impedance (BIA) scale with bare feet and recorded to nearest percentage. | Body fat percentage has also been associated with higher progesterone levels but this relationship is also not clear and may be inverse U-shaped (62,73). |
