## Supplementary material for "Quantifying Contraceptive Side-Effects: A Prospective Cohort Study of Symptom Burden, Risk Factors, and Daily Life Disruption in South-Central Ethiopia": online supplemental file 3

### S3 Appendix: Correlation plot showing correlation between different side-effects among contraceptive users after 3 months of use

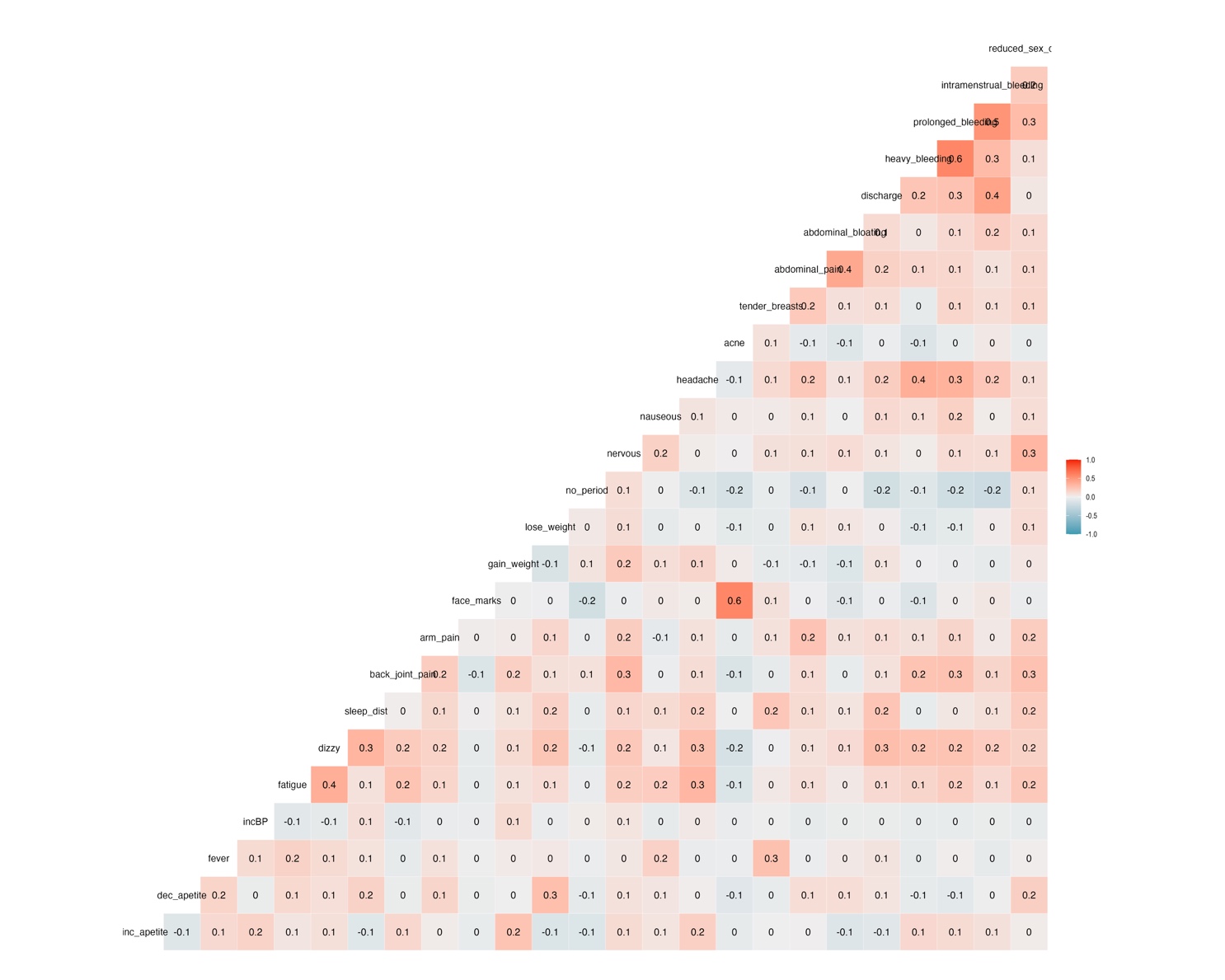
