## Supplementary material for "Quantifying Contraceptive Side-Effects: A Prospective Cohort Study of Symptom Burden, Risk Factors, and Daily Life Disruption in South-Central Ethiopia": online supplemental file 4

### S4 Appendix: Directed Acyclic Graphs and Choices of adjustment variables

**Directed Acyclic Graph showing hypothesised causal relationships between constructs included in our study**. *(No exposures or outcomes specified and including hormone levels as a hypothesised, unmeasured (oval shape) mechanistic pathway)*

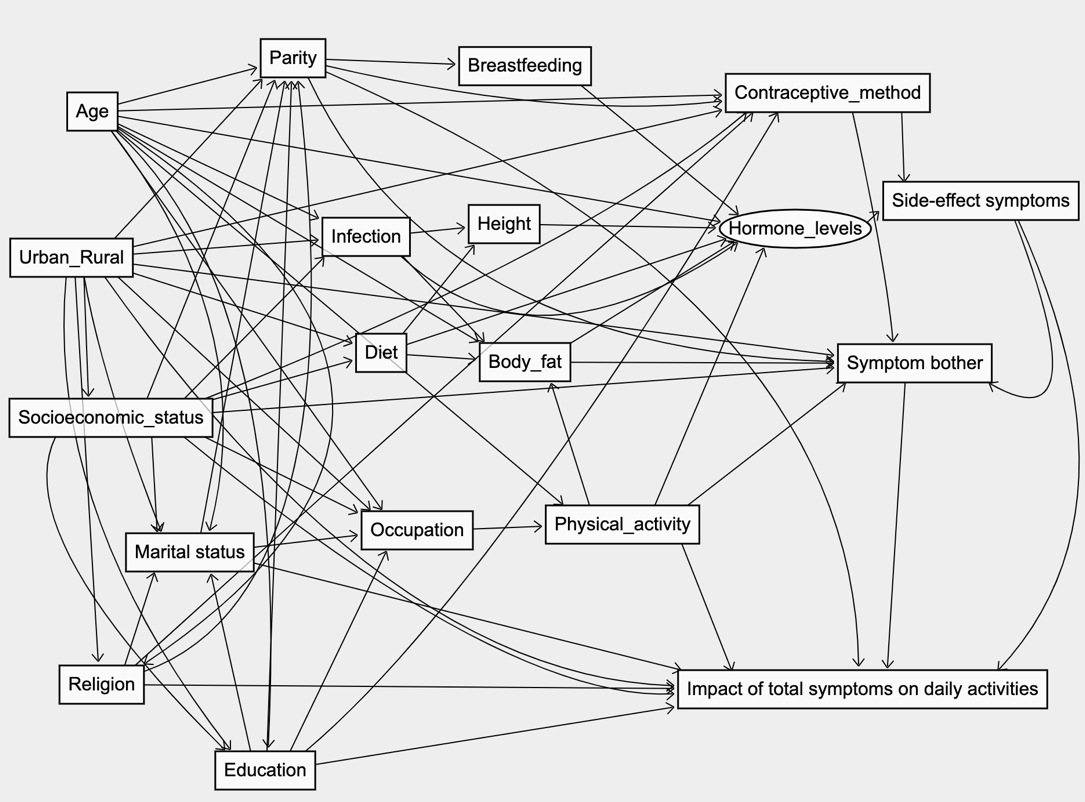

**(1) DAG for assessing change in number and severity of side-effect symptoms across contraceptive groups.**

For our first analysis, using our DAG, we identified a minimum adjustment set of age, parity, socioeconomic status, and urban/rural residence to remove any confounding ‘back doors.’ After preliminary examination of the data, we also observed high prevalence of fever at the start of the study particularly among non-users, suggesting a degree of illness upon enrolment which may bias our estimates. We hypothesise this may be driven by the high prevalence of covid in the local population around this time and disproportionate recruitment of non-users who were ill due to their contact with HEWs for treatment. Therefore, we also include as an adjustment variable whether an individual reported having a fever at baseline in interaction with the month of the survey to account for illness and recovery in our data.

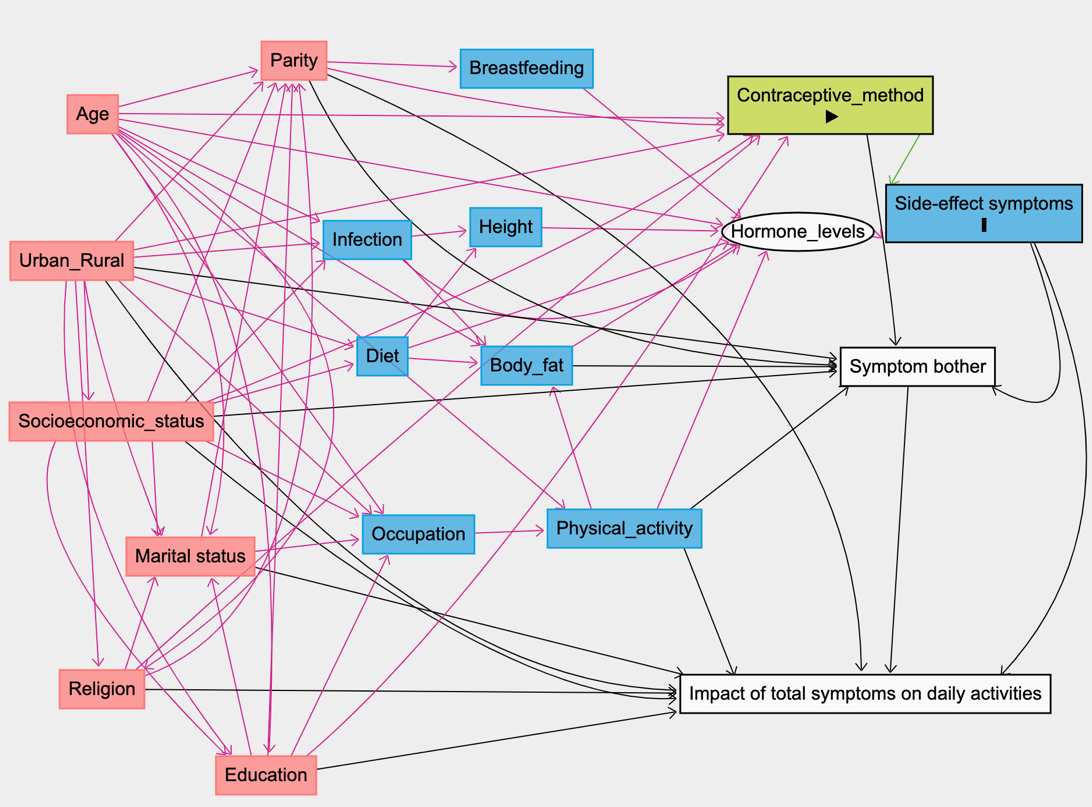

**(2) DAG for assessing risk factors for side-effects.**

When estimating the effect of physical or nutritional risk factors on side-effect symptoms, we identified a minimum adjustment set of age, urban/rural residence, education, religion, and socioeconomic status to minimise bias from confounding pathways in our estimates. Whilst adjustment variables did vary slightly depending on individual risk factors, they remained fairly consistent across risk factors and as this is an exploratory risk factor analysis, we choose to keep our adjustment factors consistent to allow comparison across estimates. Method type was also included to account for any differences between injectable and implant users.

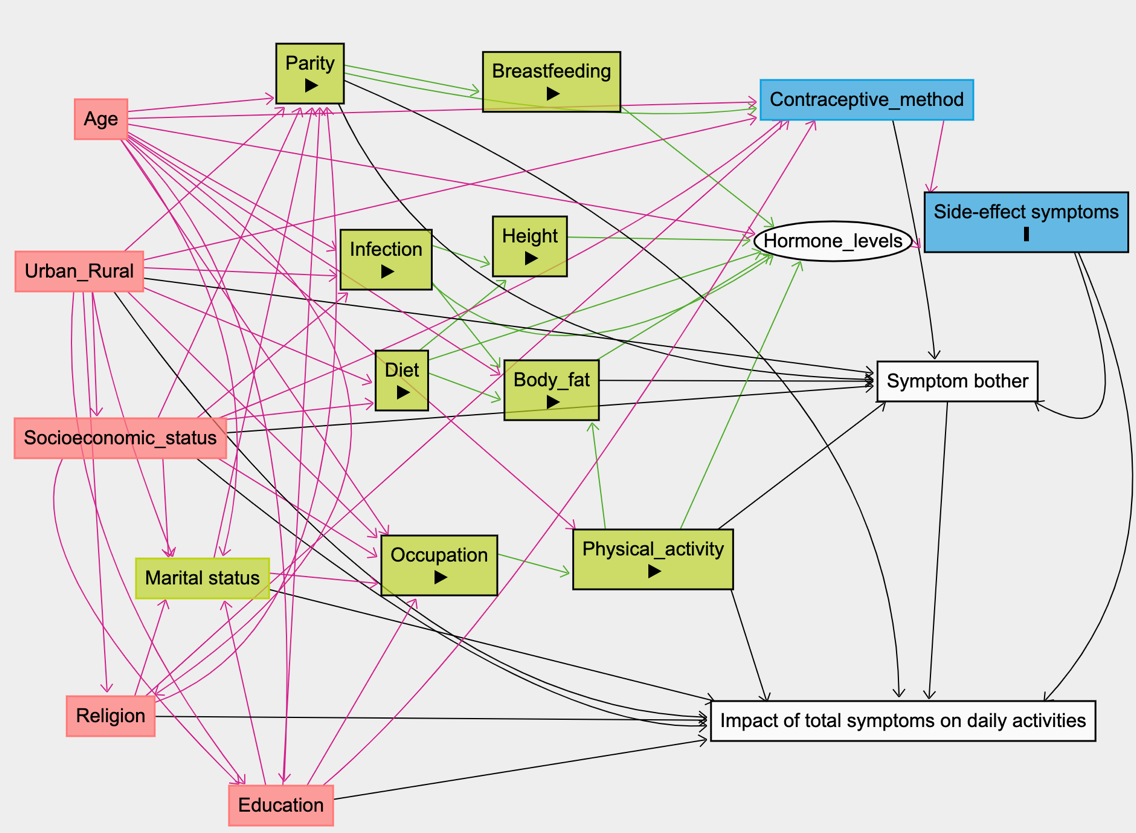

**(3) DAG for assessing impact of side-effects.**

We can see here that when estimating the effect of specific side-effect symptoms on whether an individual reported that their symptoms in the last month impacted certain daily activities, there are many potential backdoor biasing pathways. The minimum adjustment set that we chose to close of these pathways, based on this DAG, include adjusting for contraceptive group, parity, urban/rural residence, socioeconomic status, occupation, marital status, religion, and education.

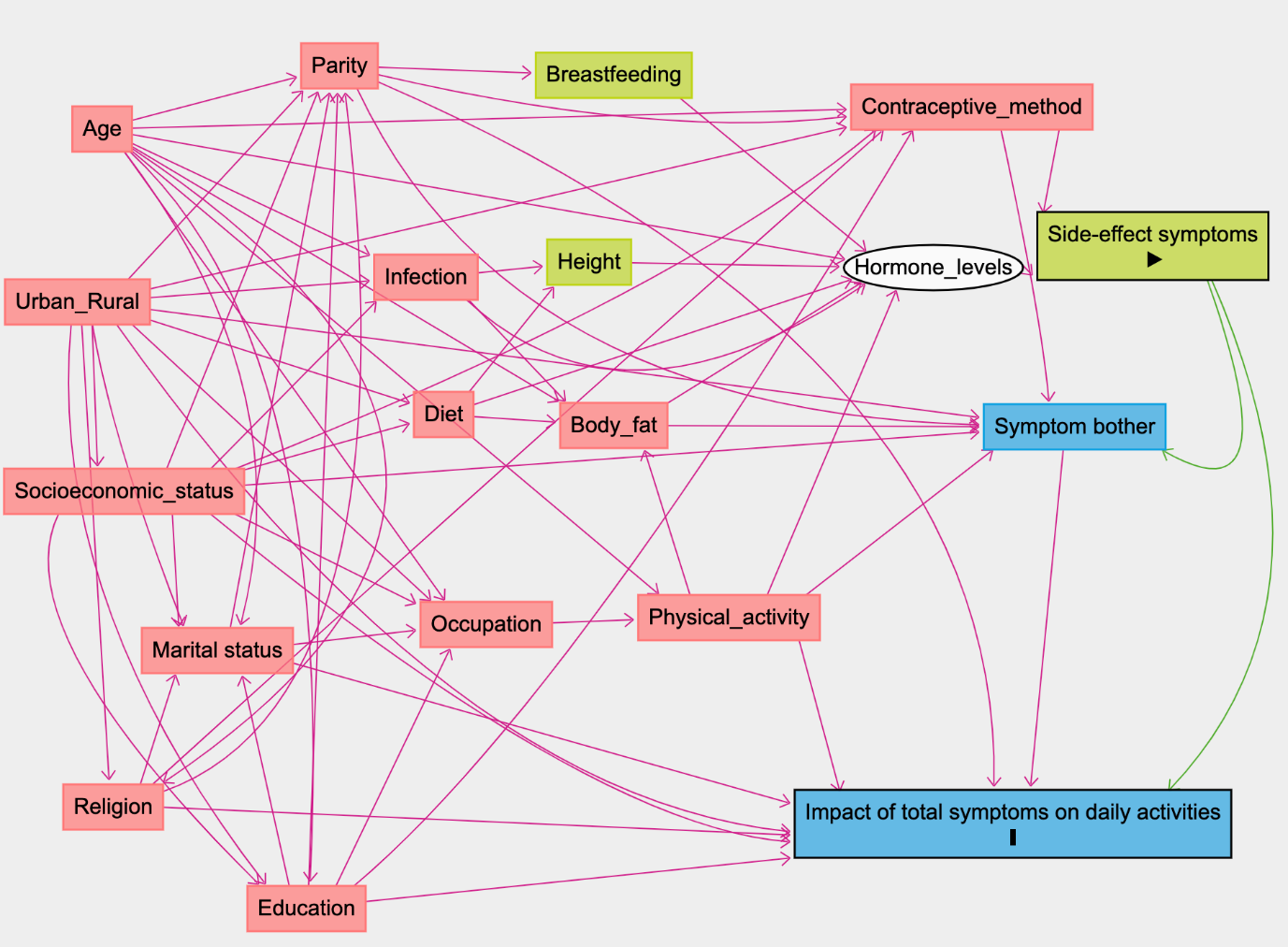
