## Supplementary material for "Quantifying Contraceptive Side-Effects: A Prospective Cohort Study of Symptom Burden, Risk Factors, and Daily Life Disruption in South-Central Ethiopia": online supplemental file 5

### S5 Appendix: Exclusion criteria used for creation of final analytical sample

368 participants were recruited into the study (Figure 1 in the paper); 152 injectable users, 100 implant, users and 116 non-users. Those who were lost to follow up or had at least one time point of side-effect measurement missing were excluded (N=37) for a complete case analysis of side-effect prevalence. Those who were unable to be reached for background data collection (N=15 total, 3 removed additionally) were also excluded. Exclusion criteria also included those who switched method or adopted a new method (N = 10) and contraceptive initiators who had start dates recorded more than a week before or after baseline symptom collection (N=40) were excluded as they violate study assumptions of consistent method usage and symptom recording upon initiation and monthly afterwards (Of the final users sample, 140 started their contraceptive on the same day as the baseline side-effects measure, 35 started up to a week after, and 3 started up to a week before). No discontinuations were observed over the study period. The final analytical sample included 278 participants; 106 injectable users, 72 implant users, and 100 non-users
