## Supplementary material for "Quantifying Contraceptive Side-Effects: A Prospective Cohort Study of Symptom Burden, Risk Factors, and Daily Life Disruption in South-Central Ethiopia": online supplemental file 6

### S6 Appendix: Characteristics of participants included and excluded from the analysis.

| **Variable** | **N** | **Overall**, N = 368*^1^* | **Excluded**, N = 90*^1^* | **Included**, N = 278*^1^* | **p-value***^2^* |
| --- | --- | --- | --- | --- | --- |
| **contraception** | 367 |  |  |  | 0.003 |
| None |  | 115 (31%) | 15 (17%) | 100 (36%) |  |
| Injectable |  | 152 (41%) | 46 (52%) | 106 (38%) |  |
| Implant |  | 100 (27%) | 28 (31%) | 72 (26%) |  |
| **# symptoms at month 0 (baseline)** | 366 | 3.0 (1.0, 5.8) | 4.0 (2.0, 6.0) | 3.0 (1.0, 5.0) | 0.078 |
| **# symptoms at month 1** | 344 | 4.00 (2.00, 6.00) | 4.00 (2.00, 6.00) | 4.00 (1.25, 6.00) | 0.2 |
| **# symptoms at month 2** | 348 | 4.00 (2.00, 6.00) | 4.00 (2.00, 7.00) | 3.00 (2.00, 6.00) | 0.039 |
| **# symptoms at month 3** | 352 | 3.00 (2.00, 5.00) | 4.00 (2.00, 6.00) | 3.00 (2.00, 5.00) | 0.004 |
| **age** | 353 | 24.0 (21.0, 27.0) | 25.0 (23.0, 27.0) | 23.0 (21.0, 27.0) | 0.013 |
| **rural/urban residence** | 353 |  |  |  | 0.028 |
| Rural |  | 162 (46%) | 26 (35%) | 136 (49%) |  |
| Urban |  | 191 (54%) | 49 (65%) | 142 (51%) |  |
| **kebele** | 353 |  |  |  | <0.001 |
| Adama City 1 |  | 48 (14%) | 12 (16%) | 36 (13%) |  |
| Adama City 14 |  | 48 (14%) | 5 (6.7%) | 43 (15%) |  |
| Adama City 4 |  | 46 (13%) | 19 (25%) | 27 (9.7%) |  |
| Adama City 9 |  | 49 (14%) | 13 (17%) | 36 (13%) |  |
| Adulala Hate |  | 48 (14%) | 18 (24%) | 30 (11%) |  |
| Awash Melkasa |  | 21 (5.9%) | 0 (0%) | 21 (7.6%) |  |
| Didimtu |  | 47 (13%) | 3 (4.0%) | 44 (16%) |  |
| Wenji Kuriftu |  | 46 (13%) | 5 (6.7%) | 41 (15%) |  |
| **highest education level attained** | 353 |  |  |  | 0.6 |
| None/Primary School |  | 128 (36%) | 27 (36%) | 101 (36%) |  |
| Secondary School |  | 118 (33%) | 22 (29%) | 96 (35%) |  |
| Higher education/technical training |  | 107 (30%) | 26 (35%) | 81 (29%) |  |
| **occupation** | 353 |  |  |  | 0.2 |
| housewife or none |  | 174 (49%) | 41 (55%) | 133 (48%) |  |
| non-physical job or student |  | 135 (38%) | 29 (39%) | 106 (38%) |  |
| physical job |  | 44 (12%) | 5 (6.7%) | 39 (14%) |  |
| **socioeconomic quintile** | 348 |  |  |  | 0.035 |
| 1 |  | 38 (11%) | 5 (6.9%) | 33 (12%) |  |
| 2 |  | 68 (20%) | 7 (9.7%) | 61 (22%) |  |
| 3 |  | 98 (28%) | 24 (33%) | 74 (27%) |  |
| 4 |  | 66 (19%) | 13 (18%) | 53 (19%) |  |
| 5 |  | 78 (22%) | 23 (32%) | 55 (20%) |  |
| **had anaemia in the last year** | 353 |  |  |  | 0.025 |
| had anaemia |  | 71 (20%) | 22 (29%) | 49 (18%) |  |
| no anaemia |  | 282 (80%) | 53 (71%) | 229 (82%) |  |
| **time since last birth** | 231 |  |  |  | 0.6 |
| 0-6 months ago |  | 55 (24%) | 14 (24%) | 41 (24%) |  |
| 13-24 months ago |  | 59 (26%) | 18 (31%) | 41 (24%) |  |
| 7-12 months ago |  | 62 (27%) | 15 (26%) | 47 (27%) |  |
| Over 24 months ago |  | 55 (24%) | 11 (19%) | 44 (25%) |  |
| **ever been pregnant** | 353 |  |  |  | 0.012 |
| Has been pregnant |  | 235 (67%) | 59 (79%) | 176 (63%) |  |
| Never been pregnant |  | 118 (33%) | 16 (21%) | 102 (37%) |  |
| **parity** | 353 |  |  |  | 0.001 |
| 0 |  | 122 (35%) | 17 (23%) | 105 (38%) |  |
| 1 |  | 125 (35%) | 23 (31%) | 102 (37%) |  |
| 2+ |  | 106 (30%) | 35 (47%) | 71 (26%) |  |
| **Breastfeeding status** | 353 |  |  |  | 0.035 |
| Currently breastfeeding |  | 146 (41%) | 39 (52%) | 107 (38%) |  |
| Not currently breastfeeding |  | 207 (59%) | 36 (48%) | 171 (62%) |  |
| **age at menarche** | 351 | 14 (14, 15) | 14 (14, 15) | 14 (14, 15) | 0.9 |
| **weight** | 353 | 52 (46, 58) | 53 (46, 61) | 51 (46, 57) | 0.2 |
| **BMI** | 353 | 20.7 (18.4, 23.1) | 21.6 (18.4, 24.2) | 20.5 (18.4, 22.8) | 0.2 |
| **body fat percentage** | 344 | 25 (18, 30) | 26 (20, 33) | 24 (18, 30) | 0.062 |
| **egg consumption** | 350 |  |  |  | 0.14 |
| More than twice a week |  | 97 (28%) | 26 (36%) | 71 (26%) |  |
| 1-2 times a week |  | 138 (39%) | 29 (40%) | 109 (39%) |  |
| Less than once a week |  | 115 (33%) | 18 (25%) | 97 (35%) |  |
| **meat consumption** | 351 |  |  |  | 0.082 |
| More than once a month |  | 163 (46%) | 41 (55%) | 122 (44%) |  |
| Less than once a month |  | 188 (54%) | 33 (45%) | 155 (56%) |  |
| **food security** | 353 |  |  |  | 0.2 |
| Food insecure |  | 98 (28%) | 16 (21%) | 82 (29%) |  |
| Food secure |  | 255 (72%) | 59 (79%) | 196 (71%) |  |
| **had a smaller meal than usual in last 4 weeks** | 353 | 88 (25%) | 15 (20%) | 73 (26%) | 0.3 |
| **had fewer meals than usual in last 4 weeks** | 353 | 57 (16%) | 9 (12%) | 48 (17%) | 0.3 |
| **had no food in the house in last 4 weeks** | 353 | 8 (2.3%) | 0 (0%) | 8 (2.9%) | 0.2 |
| **went to sleep hungry in last 4 weeks** | 353 | 12 (3.4%) | 2 (2.7%) | 10 (3.6%) | >0.9 |
| **MET minutes of activity (WHO measure)** | 348 | 2,680 (1,550, 5,580) | 2,720 (1,400, 4,280) | 2,640 (1,560, 6,240) | 0.3 |
| **work includes vigorous exercise** | 353 | 37 (10%) | 3 (4.0%) | 34 (12%) | 0.039 |
| **work includes moderate exercise** | 353 | 334 (95%) | 72 (96%) | 262 (94%) | 0.8 |
| **average time spent walking/cycling for travel per day** | 350 | 40 (30, 60) | 40 (30, 60) | 40 (30, 60) | 0.7 |
| *^1^* n (%); Median (IQR) | | | | | |
| *^2^* Pearson’s Chi-squared test; Wilcoxon rank sum test; Fisher’s exact test | | | | | |
