## Supplementary material for "Quantifying Contraceptive Side-Effects: A Prospective Cohort Study of Symptom Burden, Risk Factors, and Daily Life Disruption in South-Central Ethiopia": online supplemental file 7

### S7: Descriptive statistics across groups

*Data are n (%) or mean (SD). P values are calculated using Kruskal-Wallis rank sum test and Pearson’s Chi-squared test.*

| **Variable** | **N** | **Overall**,  N = 278*^1^* | **None**,  N = 100*^1^* | **Injectable**,  N = 106*^1^* | **Implant**,  N = 72*^1^* | **p-value***^2^* |
| --- | --- | --- | --- | --- | --- | --- |
| **Mean number of symptoms experienced** | | | | | | |
| Baseline | 278 | 3.9 (3.4) | 4.0 (3.6) | 3.8 (3.4) | 3.8 (3.2) | >0.9 |
| Month 1 | 278 | 3.8 (2.9) | 3.5 (3.1) | 3.9 (2.6) | 4.3 (3.0) | 0.2 |
| Month 2 | 278 | 3.7 (2.8) | 2.8 (2.5) | 4.1 (2.6) | 4.5 (3.3) | <0.001 |
| Month 3 | 278 | 3.6 (2.8) | 2.6 (2.3) | 4.2 (2.9) | 4.1 (3.0) | <0.001 |
| **Mean summed severity of symptoms experienced** | | | | | | |
| Baseline | 278 | 6.0 (6.0) | 6.0 (6.4) | 5.6 (6.2) | 5.7 (5.9) | >0.9 |
| Month 1 | 278 | 5.5 (5.0) | 4.7 (4.9) | 5.6 (4.5) | 6.5 (5.6) | 0.059 |
| Month 2 | 278 | 5.3 (5.0) | 3.8 (4.0) | 5.9 (4.4) | 6.7 (6.3) | <0.001 |
| Month 3 | 278 | 5.3 (5.4) | 3.6 (3.6) | 6.3 (5.7) | 6.2 (6.5) | <0.001 |
| **Age** | 278 | 23.0 (21.0, 27.0) | 22.0 (20.0, 24.2) | 25.0 (23.0, 28.0) | 23.0 (21.0, 27.0) | <0.001 |
| **Rural/urban residence** | 278 |  |  |  |  | 0.4 |
| Rural |  | 136 (49%) | 45 (45%) | 51 (48%) | 40 (56%) |  |
| Urban |  | 142 (51%) | 55 (55%) | 55 (52%) | 32 (44%) |  |
| **Highest education level attained** | 278 |  |  |  |  | <0.001 |
| None/Primary School |  | 101 (36%) | 19 (19%) | 48 (45%) | 34 (47%) |  |
| Higher education/technical training |  | 81 (29%) | 43 (43%) | 24 (23%) | 14 (19%) |  |
| Secondary School |  | 96 (35%) | 38 (38%) | 34 (32%) | 24 (33%) |  |
| **Religion** | 278 |  |  |  |  | 0.028 |
| Orthodox Christian |  | 152 (55%) | 42 (42%) | 66 (62%) | 44 (61%) |  |
| Muslim |  | 37 (13%) | 19 (19%) | 11 (10%) | 7 (9.7%) |  |
| Protestant and other |  | 89 (32%) | 39 (39%) | 29 (27%) | 21 (29%) |  |
| **Currently married** | 278 | 166 (60%) | 12 (12%) | 89 (84%) | 65 (90%) | <0.001 |
| **Socioeconomic quintile** | 276 |  |  |  |  | >0.9 |
| 1 (Lowest) |  | 33 (12%) | 11 (11%) | 12 (11%) | 10 (14%) |  |
| 2 |  | 61 (22%) | 21 (21%) | 27 (26%) | 13 (18%) |  |
| 3 |  | 74 (27%) | 26 (26%) | 25 (24%) | 23 (32%) |  |
| 4 |  | 53 (19%) | 19 (19%) | 22 (21%) | 12 (17%) |  |
| 5 (Highest) |  | 55 (20%) | 22 (22%) | 19 (18%) | 14 (19%) |  |
| Unknown |  | 2 | 1 | 1 | 0 |  |
| **Occupation** | 278 |  |  |  |  | <0.001 |
| housewife or none |  | 133 (48%) | 27 (27%) | 57 (54%) | 49 (68%) |  |
| non-physical job or student |  | 106 (38%) | 63 (63%) | 29 (27%) | 14 (19%) |  |
| physical job |  | 39 (14%) | 10 (10%) | 20 (19%) | 9 (12%) |  |
| **Work includes vigorous exercise** | 278 | 34 (12%) | 10 (10%) | 13 (12%) | 11 (15%) | 0.6 |
| **Work includes moderate exercise** | 278 | 262 (94%) | 93 (93%) | 99 (93%) | 70 (97%) | 0.5 |
| **MET minutes of activity (WHO measure)** | 273 | 2,640 (1,560, 6,240) | 2,160 (1,440, 4,320) | 3,600 (1,560, 6,720) | 3,560 (1,860, 6,640) | 0.005 |
| Unknown |  | 5 | 1 | 3 | 1 |  |
| **Average time spent walking/cycling for travel per day** | 275 | 40 (30, 60) | 55 (30, 60) | 60 (30, 60) | 40 (30, 60) | 0.7 |
| Unknown |  | 3 | 0 | 3 | 0 |  |
| **Food security** | 278 |  |  |  |  | 0.2 |
| Food secure |  | 196 (71%) | 77 (77%) | 70 (66%) | 49 (68%) |  |
| Food insecure |  | 82 (29%) | 23 (23%) | 36 (34%) | 23 (32%) |  |
| **Had a smaller meal than usual in last 4 weeks** | 278 | 73 (26%) | 18 (18%) | 35 (33%) | 20 (28%) | 0.047 |
| **Had fewer meals than usual in last 4 weeks** | 278 | 48 (17%) | 14 (14%) | 21 (20%) | 13 (18%) | 0.5 |
| **Had no food in the house in last 4 weeks** | 278 | 8 (2.9%) | 2 (2.0%) | 5 (4.7%) | 1 (1.4%) | 0.5 |
| **Went to sleep hungry in last 4 weeks** | 278 | 10 (3.6%) | 2 (2.0%) | 7 (6.6%) | 1 (1.4%) | 0.2 |
| **Egg consumption** | 277 |  |  |  |  | 0.7 |
| More than twice a week |  | 71 (26%) | 24 (24%) | 27 (25%) | 20 (28%) |  |
| 1-2 times a week |  | 109 (39%) | 45 (45%) | 38 (36%) | 26 (37%) |  |
| Less than once a week |  | 97 (35%) | 31 (31%) | 41 (39%) | 25 (35%) |  |
| Unknown |  | 1 | 0 | 0 | 1 |  |
| **Meat consumption** | 277 |  |  |  |  | 0.2 |
| More than once a month |  | 122 (44%) | 37 (37%) | 48 (46%) | 37 (51%) |  |
| Less than once a month |  | 155 (56%) | 63 (63%) | 57 (54%) | 35 (49%) |  |
| Unknown |  | 1 | 0 | 1 | 0 |  |
| **Had anaemia in the last year** | 278 |  |  |  |  | 0.012 |
| no anaemia |  | 229 (82%) | 91 (91%) | 80 (75%) | 58 (81%) |  |
| had anaemia |  | 49 (18%) | 9 (9.0%) | 26 (25%) | 14 (19%) |  |
| **Experienced any of the three symptoms of infection in last 3 months** | 278 |  |  |  |  | 0.4 |
| no symptoms of infection |  | 172 (62%) | 58 (58%) | 65 (61%) | 49 (68%) |  |
| had symptoms of infection |  | 106 (38%) | 42 (42%) | 41 (39%) | 23 (32%) |  |
| **Age at menarche** | 276 | 14 (14, 15) | 14 (13, 15) | 14 (14, 15) | 15 (14, 15) | 0.3 |
| Unknown |  | 2 | 1 | 1 | 0 |  |
| **Ever been pregnant** | 278 |  |  |  |  | <0.001 |
| Never been pregnant |  | 102 (37%) | 85 (85%) | 15 (14%) | 2 (2.8%) |  |
| Has been pregnant |  | 176 (63%) | 15 (15%) | 91 (86%) | 70 (97%) |  |
| **Parity** | 278 |  |  |  |  | <0.001 |
| 0 |  | 105 (38%) | 86 (86%) | 15 (14%) | 4 (5.6%) |  |
| 1 |  | 102 (37%) | 8 (8.0%) | 46 (43%) | 48 (67%) |  |
| 2+ |  | 71 (26%) | 6 (6.0%) | 45 (42%) | 20 (28%) |  |
| **Time since last birth** | 173 |  |  |  |  | 0.13 |
| 0-6 months ago |  | 41 (24%) | 2 (14%) | 17 (19%) | 22 (32%) |  |
| 13-24 months ago |  | 41 (24%) | 4 (29%) | 23 (25%) | 14 (21%) |  |
| 7-12 months ago |  | 47 (27%) | 2 (14%) | 24 (26%) | 21 (31%) |  |
| Over 24 months ago |  | 44 (25%) | 6 (43%) | 27 (30%) | 11 (16%) |  |
| Unknown |  | 105 | 86 | 15 | 4 |  |
| **Breastfeeding status** | 278 |  |  |  |  | <0.001 |
| Not currently breastfeeding |  | 171 (62%) | 96 (96%) | 56 (53%) | 19 (26%) |  |
| Currently breastfeeding |  | 107 (38%) | 4 (4.0%) | 50 (47%) | 53 (74%) |  |
| **Weight** | 278 | 51 (46, 57) | 50 (44, 56) | 51 (47, 56) | 54 (48, 60) | 0.010 |
| **Body fat percentage** | 269 | 24 (18, 30) | 22 (16, 28) | 24 (19, 29) | 27 (19, 32) | 0.024 |
| Unknown |  | 9 | 4 | 2 | 3 |  |
| **BMI** | 278 | 20.5 (18.4, 22.8) | 19.4 (18.0, 22.1) | 20.6 (18.5, 22.9) | 21.3 (19.6, 23.5) | 0.009 |
| *^1^* Median (IQR); n (%) | | | | | | |
| *^2^* Kruskal-Wallis rank sum test; Pearson’s Chi-squared test; Fisher’s exact test | | | | | | |
