## Supplementary material for "Quantifying Contraceptive Side-Effects: A Prospective Cohort Study of Symptom Burden, Risk Factors, and Daily Life Disruption in South-Central Ethiopia": online supplemental file 8

### S8 Appendix Figure 1: Percentage of Individuals Experiencing Symptoms Contraceptive Group (A) At Baseline

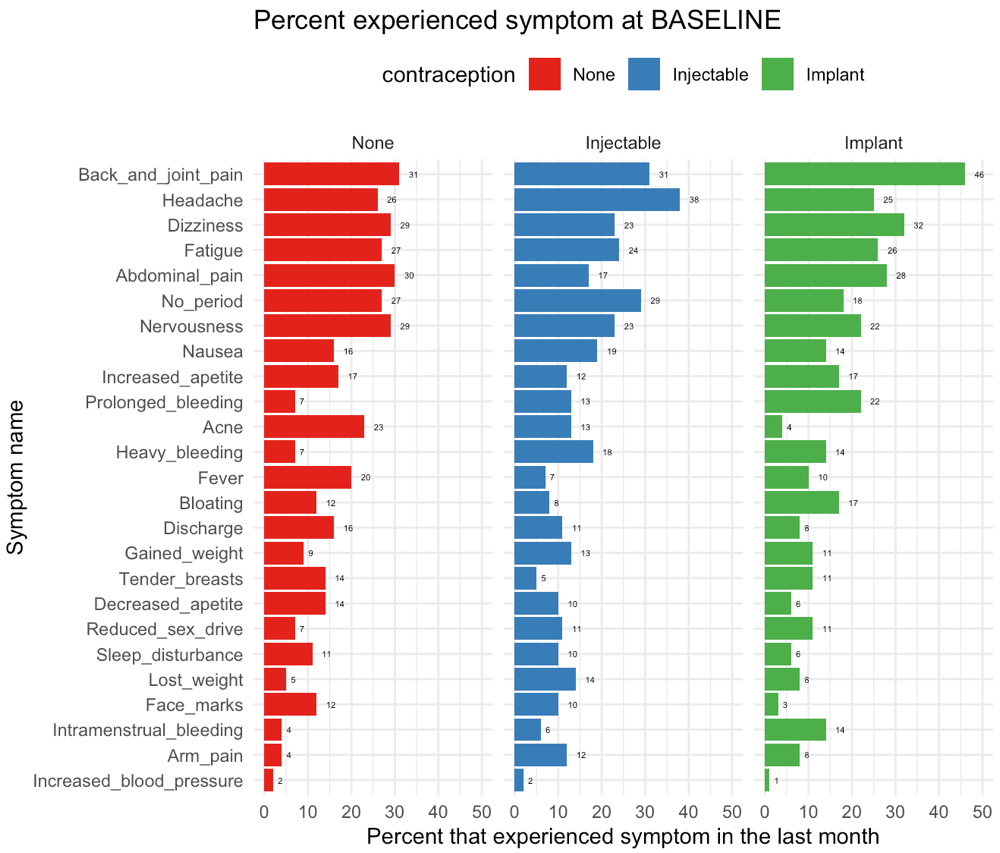

### S8 Appendix Figure 2: Percentage of Individuals Experiencing Symptoms Contraceptive Group (B) At Month 1

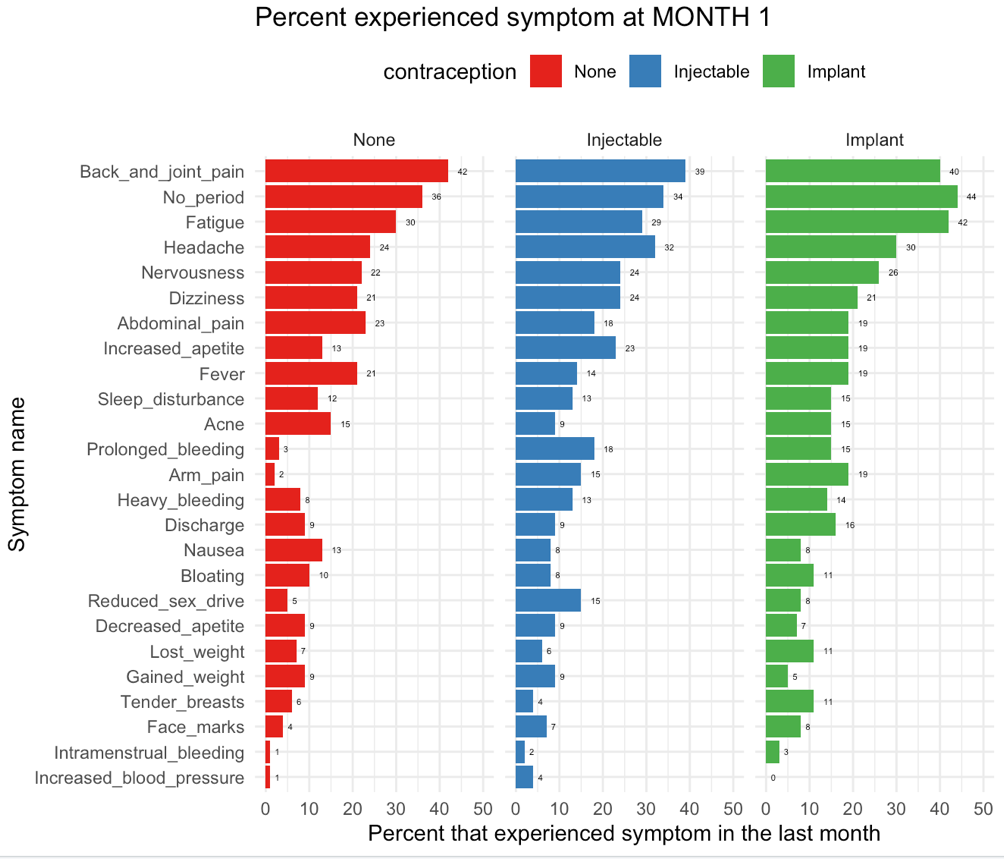

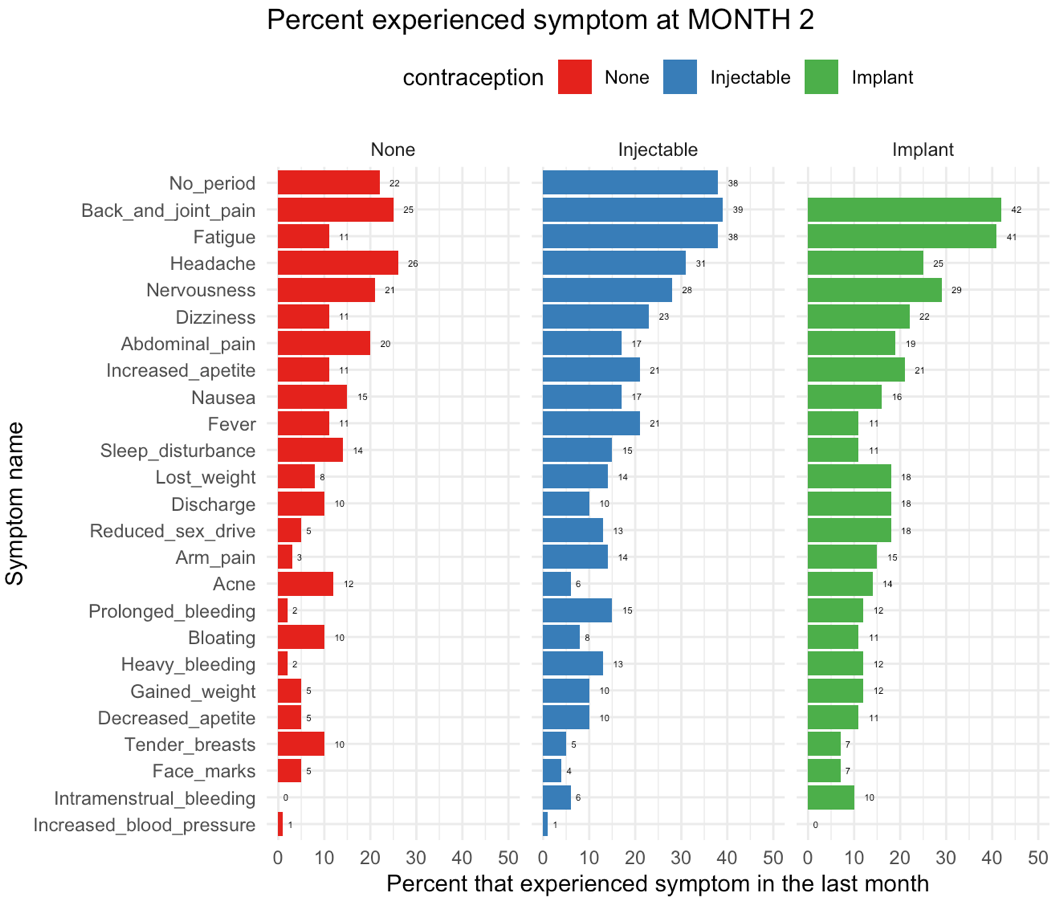

### S8 Appendix Figure 3: Percentage of Individuals Experiencing Symptoms Contraceptive Group (C) At Month 2

### S8 Appendix Figure 4: Percentage of Individuals Experiencing Symptoms Contraceptive Group (D) At Month 3

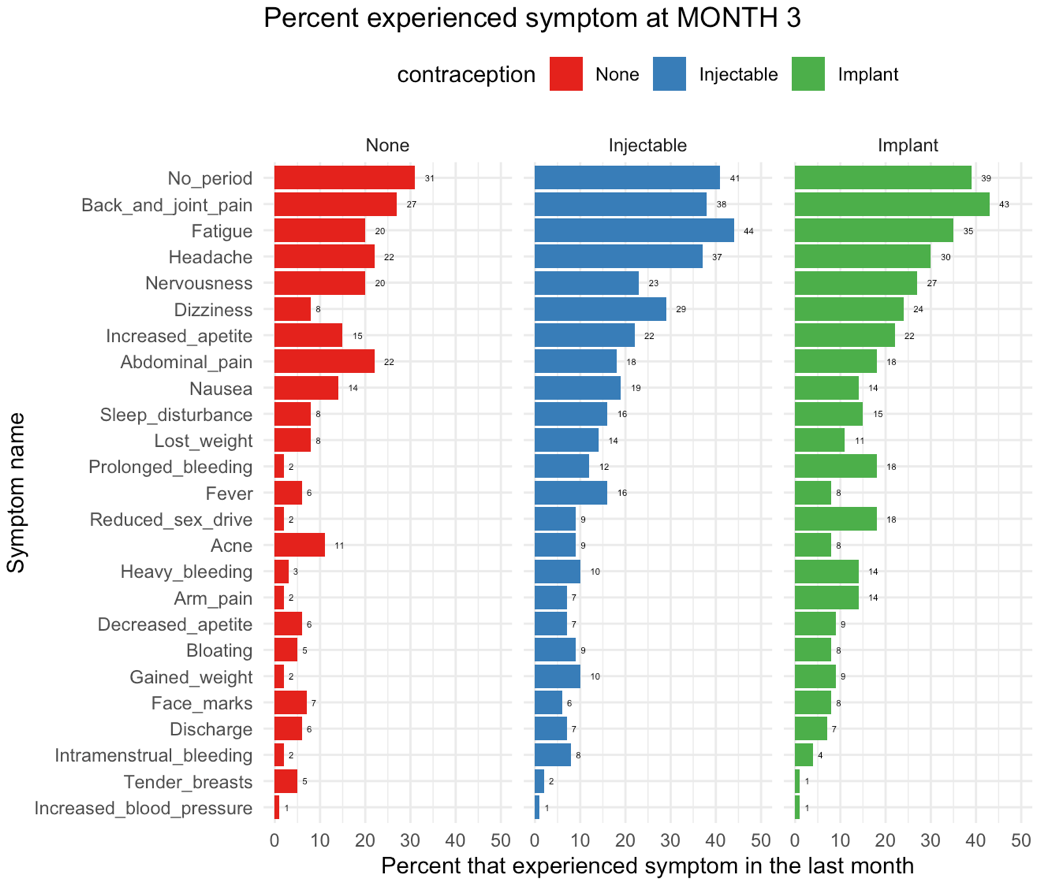

### S8 Appendix Figure 5: Percentage of Individuals Experiencing Symptoms Contraceptive Group (E) Over time

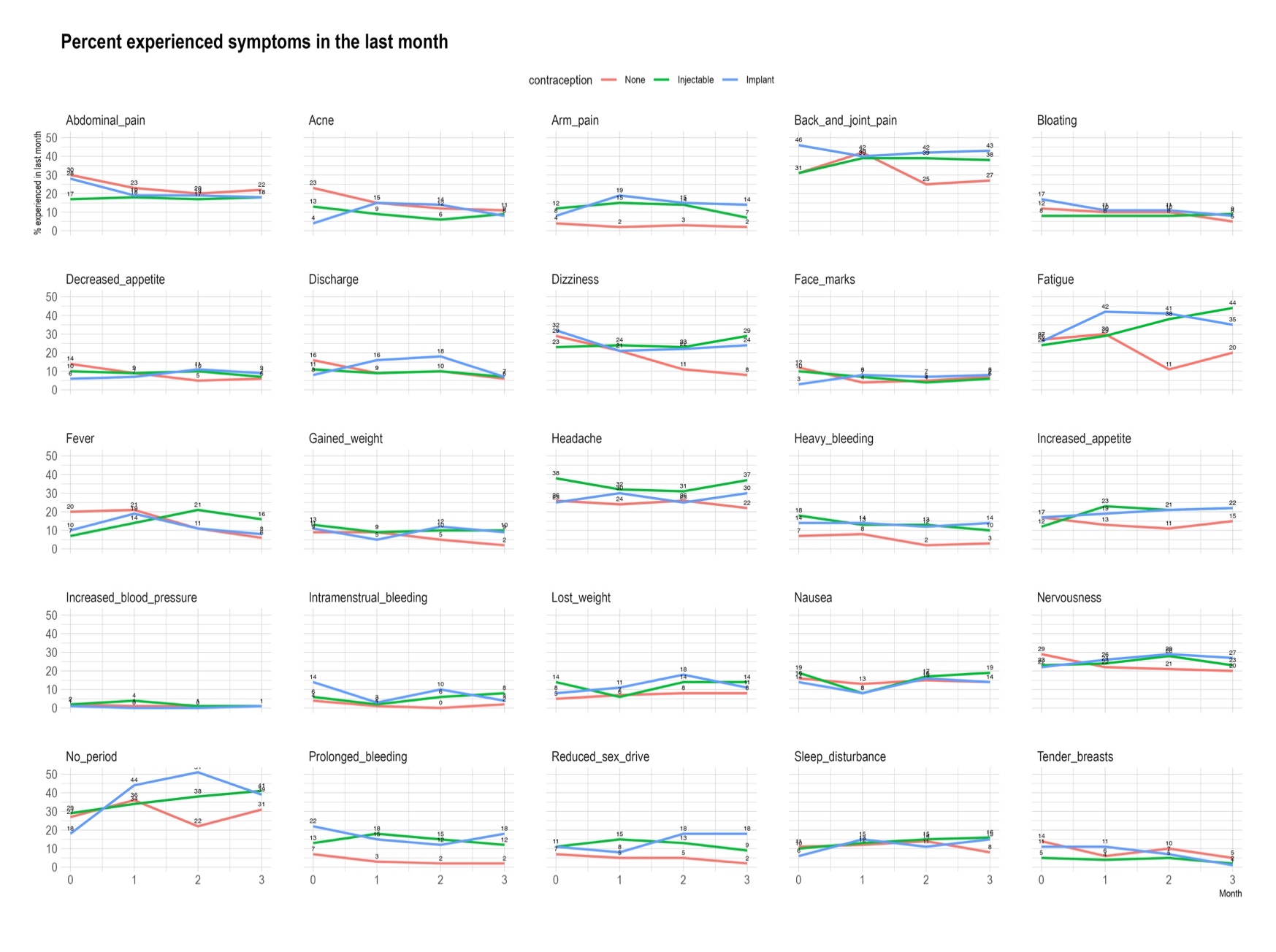
