## Supplementary material for "Quantifying Contraceptive Side-Effects: A Prospective Cohort Study of Symptom Burden, Risk Factors, and Daily Life Disruption in South-Central Ethiopia": online supplemental file 9

S9 Appendix: Evidence of association of change in each symptom over time across contraceptive groups for crude and adjusted estimates. ‘o’ represents no evidence of an association in a given month and ‘+’ or ‘–’ represent a positive or negative association respectively to p<0.05. For instance, ‘o o o +’ would represent no evidence for a change from baseline in month 1 and 2 but evidence to p<0.05 of an increase from baseline in month 3 (shown in bold). Adjustment variables include ecology, socioeconomic status, age, parity, and baseline fever in interaction with month.

|  | Crude |  |  | Adjusted |  |  |
| --- | --- | --- | --- | --- | --- | --- |
|  | None | Injectable | Implant | None | Injectable | Implant |
| Increased appetite | o o o o | **o + + +** | o o o o | o o o o | **o + + +** | o o o o |
| Decreased appetite | **o o - o** | o o o o | o o o o | o o o o | o o o o | o o o o |
| Fever | **o o o -** | **o o + +** | o o o o | o o o o | o o o o | o o o o |
| Increased blood pressure | o o o o | o o o o | o o o o | o o o o | o o o o | o o o o |
| Fatigue | **o o - o** | **o o + +** | **o + + o** | o o o o | **o o + +** | **o + + o** |
| Dizziness | **o o - -** | o o o o | o o o o | **o o o -** | o o o o | o o o o |
| Sleep disturbance | o o o o | o o o o | o o o o | o o o o | o o o o | o o o o |
| Back and joint pain | o o o o | o o o o | o o o o | o o o o | o o o o | o o o o |
| Arm pain | o o o o | o o o o | **o + o o** | o o o o | o o o o | **o + o o** |
| Face marks | **o - o o** | **o o - o** | o o o o | **o - o o** | **o o - o** | o o o o |
| Gained weight | **o o o -** | o o o o | o o o o | **o o o -** | o o o o | o o o o |
| Lost weight | o o o o | o o o o | o o o o | o o o o | o o o o | o o o o |
| Not having a period | o o o o | o o o o | **o + + +** | o o o o | o o o o | **o + + +** |
| Nervousness | o o o o | o o o o | o o o o | o o o o | o o o o | o o o o |
| Nausea | o o o o | **o - o o** | o o o o | o o o o | **o - o o** | o o o o |
| Headache | o o o o | o o o o | o o o o | o o o o | o o o o | o o o o |
| Acne | **o o - -** | o o o o | **o + + o** | o o o o | o o o o | **o + + o** |
| Breast tenderness | **o o o -** | o o o o | **o o o -** | o o o o | o o o o | **o o o -** |
| Abdominal pain | o o o o | o o o o | o o o o | o o o o | o o o o | o o o o |
| Abdominal bloating | o o o o | o o o o | o o o o | o o o o | o o o o | o o o o |
| Discharge | **o o o -** | o o o o | **o o + o** | o o o o | o o o o | **o o + o** |
| Heavy bleeding | o o o o | o o o o | o o o o | o o o o | o o o o | o o o o |
| Prolonged bleeding | o o o o | o o o o | o o o o | o o o o | o o o o | o o o o |
| Intramenstrual bleeding | o o o o | o o o o | **o - o -** | o o o o | o o o o | **o - o -** |
| Reduced sex drive or enjoyment | o o o o | o o o o | o o o o | o o o o | o o o o | o o o o |
