## Supplementary material for "Quantifying Contraceptive Side-Effects: A Prospective Cohort Study of Symptom Burden, Risk Factors, and Daily Life Disruption in South-Central Ethiopia": online supplemental file 10

S10 Appendix: Crude associations between risk factors and summed severity scores among users of contraception at month 3.

Results show incident rate ratios from individual models (adjusting only for the difference between individual baseline symptoms and mean baseline symptoms to eliminate bias by regression to the mean) for each risk factor using a Poisson distribution.

**(A) Table**

| Term | IRR | IRR lower 95% CI | IRR higher 95% CI | Statistic | p value | FDR |
| --- | --- | --- | --- | --- | --- | --- |
| **Sociodemographic** | | | | | | |
| ***Age*** | | | | | | |
| Age | **1.108** | **1.045** | **1.175** | **3.429** | **0.001** | **0.002** |
| ***Residence*** | | | | | | |
| Rural | ref |  |  |  |  |  |
| Urban | **1.813** | **1.605** | **2.048** | **9.554** | **0.000** | **0.000** |
| ***Education*** | | | | | | |
| None or Primary | ref |  |  |  |  |  |
| Higher education or technical training | **1.331** | **1.154** | **1.535** | **3.932** | **0.000** | **0.000** |
| Secondary school | 0.872 | 0.756 | 1.005 | -1.894 | 0.058 | 0.107 |
| ***Religion*** | | | | | | |
| Christian Orthodox | ref |  |  |  |  |  |
| Muslim | **1.877** | **1.599** | **2.204** | **7.688** | **0.000** | **0.000** |
| Protestant and other | 1.002 | 0.872 | 1.152 | 0.028 | 0.978 | 0.978 |
| ***Currently married*** | | | | | | |
| Currently married | 1.043 | 0.876 | 1.241 | 0.471 | 0.638 | 0.689 |
| ***Socioeconomic quintile*** | | | | | | |
| 1 | ref |  |  |  |  |  |
| 2 | 1.073 | 0.864 | 1.332 | 0.639 | 0.523 | 0.585 |
| 3 | 1.043 | 0.844 | 1.288 | 0.387 | 0.698 | 0.737 |
| 4 | 1.108 | 0.888 | 1.383 | 0.910 | 0.363 | 0.443 |
| 5 | **1.365** | **1.101** | **1.692** | **2.834** | **0.005** | **0.011** |
| **Physical Activity** | | | | | | |
| ***Occupation*** | | | | | | |
| Housewife | ref |  |  |  |  |  |
| Non-physical job or student | 0.975 | 0.841 | 1.130 | -0.343 | 0.732 | 0.755 |
| Physical job | **1.414** | **1.219** | **1.640** | **4.568** | **0.000** | **0.000** |
| ***Vigorous work activity*** | | | | | | |
| Involved vigorous activity | 0.981 | 0.823 | 1.168 | -0.219 | 0.826 | 0.843 |
| ***Moderate work activity*** | | | | | | |
| Involved moderate activity | 1.029 | 0.785 | 1.350 | 0.209 | 0.835 | 0.843 |
| ***MET weekly activity minutes*** | | | | | | |
| MET minutes | 1.020 | 0.960 | 1.083 | 0.638 | 0.524 | 0.585 |
| ***Travel activity average minutes*** | | | | | | |
| Travel activity time | 0.853 | 0.733 | 0.993 | -2.053 | 0.040 | 0.084 |
| Travel activity time squared | 1.155 | 0.997 | 1.338 | 1.920 | 0.055 | 0.107 |
| **Nutritional stress** | | | | | | |
| ***Food insecurity*** | | | | | | |
| Food insecure | 1.026 | 0.905 | 1.162 | 0.399 | 0.690 | 0.736 |
| ***Had smaller meal*** | | | | | | |
| Had smaller meal | 1.061 | 0.935 | 1.205 | 0.917 | 0.359 | 0.443 |
| ***Had fewer meals*** | | | | | | |
| Had fewer meal | **1.226** | **1.065** | **1.410** | **2.844** | **0.004** | **0.011** |
| ***Had no food in house*** | | | | | | |
| Had no food in the house | 0.821 | 0.574 | 1.173 | -1.085 | 0.278 | 0.417 |
| ***Went to sleep hungry*** | | | | | | |
| Went to sleep hungry | **1.496** | **1.179** | **1.897** | **3.317** | **0.001** | **0.002** |
| ***Egg consumption*** | | | | | | |
| More than once a week | ref |  |  |  |  |  |
| 1 to 2 times a week | 1.071 | 0.918 | 1.249 | 0.871 | 0.384 | 0.459 |
| Less than once a week | 1.147 | 0.986 | 1.333 | 1.778 | 0.075 | 0.136 |
| ***Meat consumption*** | | | | | | |
| More than once a month | ref |  |  |  |  |  |
| Less than once a month | **0.802** | **0.713** | **0.902** | **-3.685** | **0.000** | **0.001** |
| ***Anaemia in last year*** | | | | | | |
| Reported having anaemia | 1.034 | 0.900 | 1.188 | 0.470 | 0.639 | 0.689 |
| **Health and Reproduction** | | | | | | |
| ***Any recent infection symptoms*** | | | | | | |
| Had symptoms of infection in last 3 months | 1.125 | 0.997 | 1.269 | 1.915 | 0.055 | 0.107 |
| ***Age at menarche*** | | | | | | |
| Age at menarche | 0.936 | 0.882 | 0.993 | -2.195 | 0.028 | 0.061 |
| ***Ever been pregnant*** | | | | | | |
| Been pregnant | **1.297** | **1.039** | **1.619** | **2.302** | **0.021** | **0.049** |
| ***Parity*** | | | | | | |
| 0 | ref |  |  |  |  |  |
| 1 | **1.425** | **1.145** | **1.774** | **3.170** | **0.002** | **0.004** |
| 2 | 1.223 | 0.975 | 1.534 | 1.738 | 0.082 | 0.146 |
| ***Time since last birth*** | | | | | | |
| 0-6 months ago | ref |  |  |  |  |  |
| 13 to 24 months ago | **1.556** | **1.275** | **1.899** | **4.348** | **0.000** | **0.000** |
| 7 to 12 months ago | **1.846** | **1.530** | **2.226** | **6.410** | **0.000** | **0.000** |
| Over 24 months ago | **1.788** | **1.475** | **2.169** | **5.904** | **0.000** | **0.000** |
| ***Breastfeeding status*** | | | | | | |
| Currently breastfeeding | 0.892 | 0.793 | 1.004 | -1.901 | 0.057 | 0.107 |
| **Anthropometrics** | | | | | | |
| ***Weight*** | | | | | | |
| Weight | 0.974 | 0.918 | 1.034 | -0.862 | 0.389 | 0.459 |
| ***Body fat*** | | | | | | |
| Body fat | 1.324 | 0.999 | 1.756 | 1.952 | 0.051 | 0.104 |
| Body fat squared | 0.728 | 0.548 | 0.968 | -2.184 | 0.029 | 0.062 |
| ***BMI*** | | | | | | |
| BMI | 0.982 | 0.925 | 1.041 | -0.615 | 0.538 | 0.594 |

(B) Forest plot

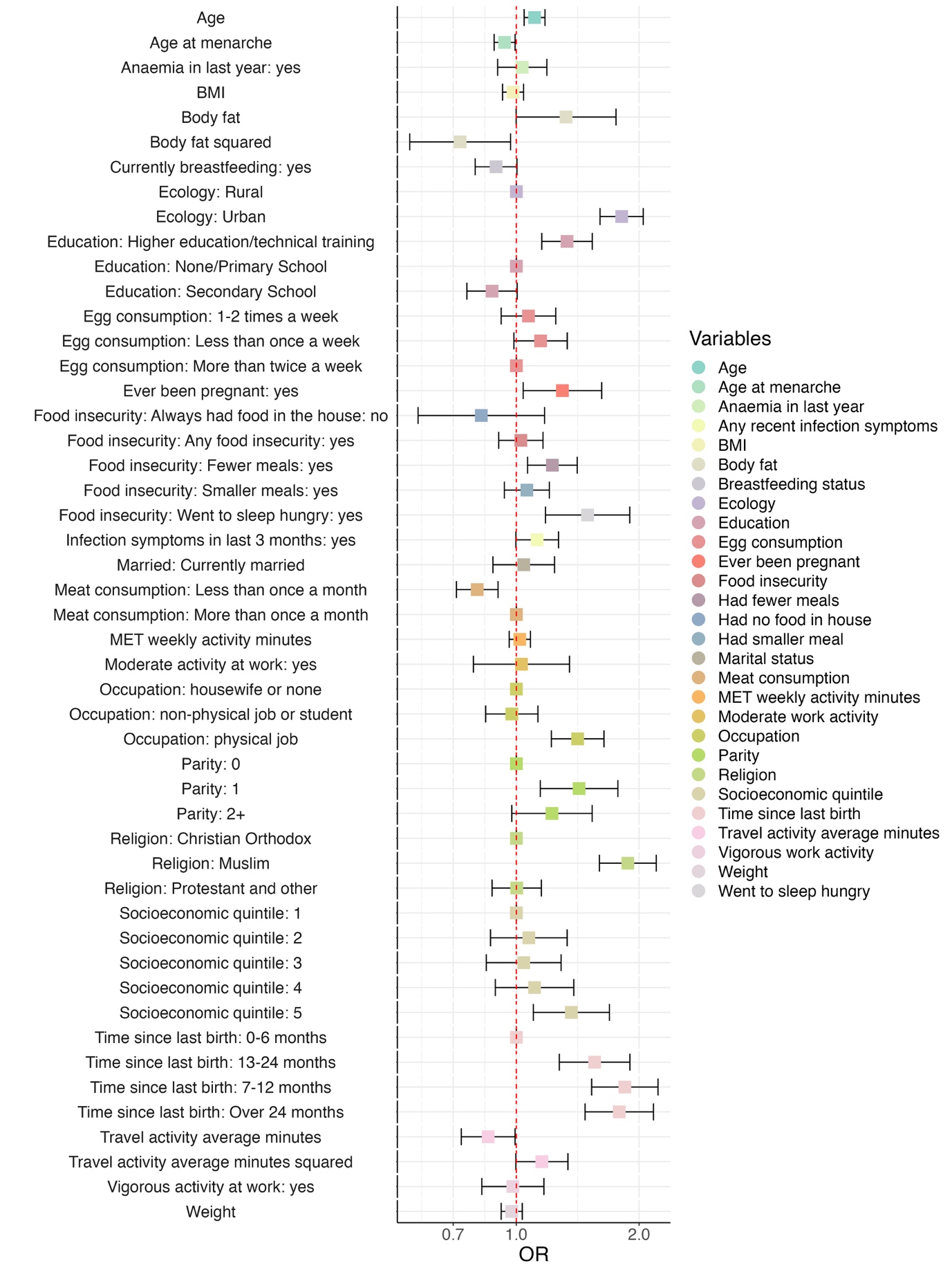
