## Supplementary material for "Quantifying Contraceptive Side-Effects: A Prospective Cohort Study of Symptom Burden, Risk Factors, and Daily Life Disruption in South-Central Ethiopia": online supplemental file 11

S11 Appendix: Adjusted associations between risk factors and summed severity scores among users of contraception at month 3.

Results show incident rate ratios from minimally-adjusted individual models for each risk factor using a Poisson distribution. Adjustment variables included age, urban/rural residence, socioeconomic status, and the difference between individual baseline symptoms and mean baseline symptoms.

| Term | IRR | IRR lower 95% CI | IRR higher 95% CI | Statistic | p value | FDR |
| --- | --- | --- | --- | --- | --- | --- |
| **Sociodemographic** | | | | | | |
| ***Currently married*** | | | | | | |
| Currently married | 1.029 | 0.855 | 1.238 | 0.303 | 0.762 | 0.803 |
| **Physical Activity** | | | | | | |
| ***Occupation*** | | | | | | |
| Housewife | ref |  |  |  |  |  |
| Non-physical job or student | **0.752** | **0.633** | **0.892** | **-3.263** | **0.001** | **0.006** |
| Physical job | **1.377** | **1.167** | **1.626** | **3.779** | **0.000** | **0.001** |
| ***Vigorous work activity*** | | | | | | |
| Involved vigorous activity | 1.244 | 1.022 | 1.515 | 2.172 | 0.030 | 0.085 |
| ***Moderate work activity*** | | | | | | |
| Involved moderate activity | 1.332 | 0.993 | 1.787 | 1.913 | 0.056 | 0.138 |
| ***MET weekly activity minutes*** | | | | | | |
| MET minutes | 1.075 | 1.004 | 1.151 | 2.080 | 0.038 | 0.099 |
| ***Travel activity average minutes*** | | | | | | |
| Travel activity time | 1.079 | 0.907 | 1.284 | 0.861 | 0.389 | 0.493 |
| Travel activity time squared | 0.972 | 0.831 | 1.137 | -0.360 | 0.719 | 0.767 |
| **Nutritional stress** | | | | | | |
| ***Food insecurity*** | | | | | | |
| Food insecure | 1.094 | 0.942 | 1.270 | 1.175 | 0.240 | 0.365 |
| ***Had smaller meal*** | | | | | | |
| Had smaller meal | 1.098 | 0.945 | 1.276 | 1.220 | 0.223 | 0.356 |
| ***Had fewer meals*** | | | | | | |
| Had fewer meal | **1.342** | **1.140** | **1.578** | **3.543** | **0.000** | **0.002** |
| ***Had no food in house*** | | | | | | |
| Had no food in the house | 0.884 | 0.604 | 1.294 | -0.632 | 0.527 | 0.607 |
| ***Went to sleep hungry*** | | | | | | |
| Went to sleep hungry | **1.474** | **1.138** | **1.910** | **2.935** | **0.003** | **0.015** |
| ***Egg consumption*** | | | | | | |
| More than once a week | ref |  |  |  |  |  |
| 1 to 2 times a week | 1.184 | 1.002 | 1.398 | 1.980 | 0.048 | 0.123 |
| Less than once a week | **1.318** | **1.103** | **1.574** | **3.043** | **0.002** | **0.011** |
| ***Meat consumption*** | | | | | | |
| More than once a month | ref |  |  |  |  |  |
| Less than once a month | **0.838** | **0.738** | **0.952** | **-2.717** | **0.007** | **0.028** |
| ***Anaemia in last year*** | | | | | | |
| Reported having anaemia | 0.945 | 0.816 | 1.096 | -0.746 | 0.456 | 0.539 |
| **Health and Reproduction** | | | | | | |
| ***Any recent infection symptoms*** | | | | | | |
| Had symptoms of infection in last 3 months | **1.273** | **1.118** | **1.449** | **3.653** | **0.000** | **0.002** |
| ***Age at menarche*** | | | | | | |
| Age at menarche | 0.934 | 0.877 | 0.995 | -2.100 | 0.036 | 0.098 |
| ***Ever been pregnant*** | | | | | | |
| Been pregnant | **1.347** | **1.067** | **1.699** | **2.510** | **0.012** | **0.042** |
| ***Parity*** | | | | | | |
| 0 | ref |  |  |  |  |  |
| 1 | **1.524** | **1.218** | **1.907** | **3.685** | **0.000** | **0.001** |
| 2 | 1.154 | 0.899 | 1.480 | 1.126 | 0.260 | 0.374 |
| ***Time since last birth*** | | | | | | |
| 0 to 6 months ago | ref |  |  |  |  |  |
| 13 to 24 months ago | 1.263 | 1.020 | 1.565 | 2.140 | 0.032 | 0.090 |
| 7 to 12 months ago | **1.731** | **1.428** | **2.099** | **5.578** | **0.000** | **0.000** |
| Over 24 months ago | **1.629** | **1.318** | **2.014** | **4.516** | **0.000** | **0.000** |
| ***Breastfeeding status*** | | | | | | |
| Currently breastfeeding | 1.011 | 0.885 | 1.155 | 0.159 | 0.874 | 0.901 |
| **Anthropometrics** | | | | | | |
| ***Weight*** | | | | | | |
| Weight | 0.939 | 0.877 | 1.006 | -1.793 | 0.073 | 0.159 |
| ***Body fat*** | | | | | | |
| Body fat | **1.624** | **1.217** | **2.166** | **3.292** | **0.001** | **0.006** |
| Body fat squared | **0.563** | **0.418** | **0.759** | **-3.775** | **0.000** | **0.001** |
| ***BMI*** | | | | | | |
| BMI | 0.951 | 0.886 | 1.021 | -1.388 | 0.165 | 0.282 |
