## Supplementary material for "Quantifying Contraceptive Side-Effects: A Prospective Cohort Study of Symptom Burden, Risk Factors, and Daily Life Disruption in South-Central Ethiopia": online supplemental file 12

**S12 Appendix Table: Adjusted association between risk factors and summed severity scores among users of contraception at month 2.**

Results show incident rate ratios from minimally-adjusted individual models for each risk factor using a Poisson distribution. Adjustment variables included age, urban/rural residence, socioeconomic status, and the difference between individual baseline symptoms and mean baseline symptoms.

| Term | IRR | IRR lower 95% CI | IRR higher 95% CI | Statistic | p value | FDR |
| --- | --- | --- | --- | --- | --- | --- |
| **Sociodemographic** | | | | | | |
| ***Currently married*** | | | | | | |
| Currently married | 0.909 | 0.757 | 1.092 | -1.016 | 0.310 | 0.430 |
| **Physical Activity** | | | | | | |
| ***Occupation*** | | | | | | |
| Housewife | ref |  |  |  |  |  |
| Non-physical job or student | **0.734** | **0.616** | **0.875** | **-3.451** | **0.001** | **0.002** |
| Physical job | 1.195 | 1.008 | 1.416 | 2.053 | 0.040 | 0.079 |
| ***Vigorous work activity*** | | | | | | |
| Involved vigorous activity | **1.812** | **1.513** | **2.171** | **6.454** | **0.000** | **0.000** |
| ***Moderate work activity*** | | | | | | |
| Involved moderate activity | 1.088 | 0.815 | 1.452 | 0.573 | 0.567 | 0.698 |
| ***MET weekly activity minutes*** | | | | | | |
| MET minutes | **1.206** | **1.132** | **1.285** | **5.810** | **0.000** | **0.000** |
| ***Travel activity average minutes*** | | | | | | |
| Travel activity time | 0.961 | 0.808 | 1.144 | -0.447 | 0.655 | 0.788 |
| Travel activity time squared | 1.120 | 0.960 | 1.306 | 1.440 | 0.150 | 0.229 |
| **Nutritional stress** | | | | | | |
| ***Food insecurity*** | | | | | | |
| Food insecure | **1.198** | **1.032** | **1.390** | **2.377** | **0.017** | **0.037** |
| ***Had smaller meal*** | | | | | | |
| Had smaller meal | 1.157 | 0.996 | 1.344 | 1.904 | 0.057 | 0.099 |
| ***Had fewer meals*** | | | | | | |
| Had fewer meal | **1.271** | **1.078** | **1.498** | **2.853** | **0.004** | **0.011** |
| ***Had no food in house*** | | | | | | |
| Had no food in the house | **1.468** | **1.060** | **2.032** | **2.314** | **0.021** | **0.043** |
| ***Went to sleep hungry*** | | | | | | |
| Went to sleep hungry | **1.575** | **1.211** | **2.050** | **3.382** | **0.001** | **0.002** |
| ***Egg consumption*** | | | | | | |
| More than once a week | ref |  |  |  |  |  |
| 1 to 2 times a week | 1.067 | 0.898 | 1.267 | 0.733 | 0.464 | 0.587 |
| Less than once a week | **1.436** | **1.200** | **1.717** | **3.955** | **0.000** | **0.000** |
| ***Meat consumption*** | | | | | | |
| More than once a month | ref |  |  |  |  |  |
| Less than once a month | 1.026 | 0.902 | 1.167 | 0.393 | 0.694 | 0.827 |
| ***Anaemia in last year*** | | | | | | |
| Reported having anaemia | **1.349** | **1.173** | **1.551** | **4.200** | **0.000** | **0.000** |
| **Health and Reproduction** | | | | | | |
| ***Any recent infection symptoms*** | | | | | | |
| Had symptoms of infection in last 3 months | 1.059 | 0.926 | 1.211 | 0.841 | 0.400 | 0.517 |
| ***Age at menarche*** | | | | | | |
| Age at menarche | 0.940 | 0.882 | 1.002 | -1.905 | 0.057 | 0.099 |
| ***Ever been pregnant*** | | | | | | |
| Been pregnant | 1.100 | 0.884 | 1.369 | 0.855 | 0.393 | 0.511 |
| ***Parity*** | | | | | | |
| 0 | ref |  |  |  |  |  |
| 1 | 1.052 | 0.856 | 1.293 | 0.484 | 0.628 | 0.762 |
| 2 | 1.095 | 0.868 | 1.380 | 0.765 | 0.444 | 0.565 |
| ***Time since last birth*** | | | | | | |
| 0 to 6 months | ref |  |  |  |  |  |
| 13 to 24 months ago | **1.406** | **1.142** | **1.731** | **3.216** | **0.001** | **0.004** |
| 7 to 12 months ago | **1.427** | **1.178** | **1.729** | **3.638** | **0.000** | **0.001** |
| Over 24 months ago | **1.407** | **1.134** | **1.745** | **3.108** | **0.002** | **0.006** |
| ***Breastfeeding status*** | | | | | | |
| Currently breastfeeding | 1.057 | 0.923 | 1.211 | 0.807 | 0.420 | 0.539 |
| **Anthropometrics** | | | | | | |
| ***Weight*** | | | | | | |
| Weight | 1.025 | 0.959 | 1.097 | 0.730 | 0.465 | 0.588 |
| ***Body fat*** | | | | | | |
| Body fat | 1.185 | 0.904 | 1.554 | 1.229 | 0.219 | 0.326 |
| Body fat squared | 0.861 | 0.652 | 1.136 | -1.058 | 0.290 | 0.404 |
| ***BMI*** | | | | | | |
| BMI | 1.045 | 0.974 | 1.121 | 1.218 | 0.223 | 0.330 |
