## Supplementary material for "Quantifying Contraceptive Side-Effects: A Prospective Cohort Study of Symptom Burden, Risk Factors, and Daily Life Disruption in South-Central Ethiopia": online supplemental file 13

**S13 Appendix Table: Adjusted association between risk factors and summed severity scores among non-users at month 3.**

Results show incident rate ratios from minimally-adjusted individual models for each risk factor using a Poisson distribution. Adjustment variables included age, urban/rural residence, socioeconomic status, and the difference between individual baseline symptoms and mean baseline symptoms.

| Term | IRR | IRR lower 95% CI | IRR higher 95% CI | Statistic | p value | FDR |
| --- | --- | --- | --- | --- | --- | --- |
| **Sociodemographic** | | | | | | |
| ***Currently married*** | | | | | | |
| Currently married | 0.931 | 0.615 | 1.409 | -0.339 | 0.735 | 0.801 |
| **Physical Activity** | | | | | | |
| ***Occupation*** | | | | | | |
| Housewife | ref |  |  |  |  |  |
| Non-physical job or student | 0.997 | 0.749 | 1.326 | -0.022 | 0.982 | 1.000 |
| Physical job | 1.370 | 0.911 | 2.059 | 1.512 | 0.131 | 0.197 |
| ***Vigorous work activity*** | | | | | | |
| Involved vigorous activity | 1.286 | 0.883 | 1.874 | 1.313 | 0.189 | 0.262 |
| ***Moderate work activity*** | | | | | | |
| Involved moderate activity | **0.456** | **0.313** | **0.664** | **-4.097** | **0.000** | **0.003** |
| ***MET weekly activity minutes*** | | | | | | |
| MET minutes | **1.207** | **1.074** | **1.356** | **3.164** | **0.002** | **0.010** |
| ***Travel activity average minutes*** | | | | | | |
| Travel activity time | 1.375 | 0.930 | 2.033 | 1.594 | 0.111 | 0.176 |
| Travel activity time squared | **0.549** | **0.353** | **0.854** | **-2.662** | **0.008** | **0.025** |
| **Nutritional stress** | | | | | | |
| ***Food insecurity*** | | | | | | |
| Food insecure | 0.847 | 0.623 | 1.153 | -1.054 | 0.292 | 0.361 |
| ***Had smaller meal*** | | | | | | |
| Had smaller meal | 0.774 | 0.551 | 1.087 | -1.478 | 0.139 | 0.209 |
| ***Had fewer meals*** | | | | | | |
| Had fewer meal | 0.862 | 0.594 | 1.251 | -0.781 | 0.435 | 0.518 |
| ***Had no food in house*** | | | | | | |
| Had no food in the house | 1.367 | 0.639 | 2.921 | 0.806 | 0.420 | 0.503 |
| ***Went to sleep hungry*** | | | | | | |
| Went to sleep hungry | 1.046 | 0.482 | 2.271 | 0.113 | 0.910 | 0.955 |
| ***Egg consumption*** | | | | | | |
| More than once a week | ref |  |  |  |  |  |
| 1 to 2 times a week | 0.854 | 0.651 | 1.119 | -1.145 | 0.252 | 0.329 |
| Less than once a week | 0.853 | 0.616 | 1.180 | -0.961 | 0.337 | 0.412 |
| ***Meat consumption*** | | | | | | |
| More than once a month | ref |  |  |  |  |  |
| Less than once a month | 0.934 | 0.713 | 1.224 | -0.497 | 0.619 | 0.711 |
| ***Anaemia in last year*** | | | | | | |
| Reported having anaemia | 0.756 | 0.476 | 1.200 | -1.185 | 0.236 | 0.313 |
| **Health and Reproduction** | | | | | | |
| ***Any recent infection symptoms*** | | | | | | |
| Had symptoms of infection in last 3 months | **0.680** | **0.531** | **0.871** | **-3.055** | **0.002** | **0.011** |
| ***Age at menarche*** | | | | | | |
| Age at menarche | **0.830** | **0.738** | **0.933** | **-3.126** | **0.002** | **0.010** |
| ***Ever been pregnant*** | | | | | | |
| Been pregnant | **0.629** | **0.423** | **0.936** | **-2.285** | **0.022** | **0.043** |
| ***Parity*** | | | | | | |
| 0 | ref |  |  |  |  |  |
| 1 | 0.720 | 0.460 | 1.127 | -1.435 | 0.151 | 0.224 |
| 2 | **0.429** | **0.228** | **0.806** | **-2.630** | **0.009** | **0.027** |
| ***Time since last birth*** | | | | | | |
| 0 to 6 months | ref |  |  |  |  |  |
| 13 to 24 months ago | 905.302 | 0.000 | Inf | 0.000 | 1.000 | 1.000 |
| 7 to 12 months ago | 5517609625.853 | 0.000 | Inf | 0.000 | 1.000 | 1.000 |
| Over 24 months ago | 2.840 | 0.000 | Inf | 0.000 | 1.000 | 1.000 |
| ***Breastfeeding status*** | | | | | | |
| Currently breastfeeding | 0.853 | 0.441 | 1.652 | -0.470 | 0.638 | 0.728 |
| **Anthropometrics** | | | | | | |
| ***Weight*** | | | | | | |
| Weight | 0.989 | 0.878 | 1.114 | -0.183 | 0.855 | 0.904 |
| ***Body fat*** | | | | | | |
| Body fat | **0.570** | **0.351** | **0.927** | **-2.267** | **0.023** | **0.045** |
| Body fat squared | **1.832** | **1.119** | **3.001** | **2.407** | **0.016** | **0.033** |
| ***BMI*** | | | | | | |
| BMI | 1.001 | 0.891 | 1.123 | 0.011 | 0.991 | 1.000 |
