## Supplementary material for "Quantifying Contraceptive Side-Effects: A Prospective Cohort Study of Symptom Burden, Risk Factors, and Daily Life Disruption in South-Central Ethiopia": online supplemental file 14

**S14 Appendix Table 1: Percent of surveys reporting a negative impact of symptoms experienced in the last month on 6 different daily activities at month 3 split by contraceptive group at all time points.**

N represents the number of surveys where participants undertook that activity (Total N = 278).

| Impact of symptoms by contraceptive group after 3 months | | | | | | |
| --- | --- | --- | --- | --- | --- | --- |
| **Variable** | **N** | **Overall**, N = 278*^1^* | **None**, N = 96*^1^* | **Injectable**, N = 108*^1^* | **Implant**, N = 74*^1^* | **p-value***^2^* |
| **Impacted employment** | 174 | 63 (36%) | 21 (36%) | 21 (30%) | 21 (47%) | 0.2 |
| **Impacted education** | 121 | 62 (51%) | 14 (23%) | 25 (81%) | 23 (79%) | <0.001 |
| **Impacted daily chores** | 276 | 37 (13%) | 9 (9.6%) | 15 (14%) | 13 (18%) | 0.3 |
| **Impacted ability to care for children** | 186 | 17 (9.1%) | 0 (0%) | 10 (10%) | 7 (10%) | 0.6 |
| **Impacted harmonious relationship** | 173 | 20 (12%) | 2 (14%) | 11 (12%) | 7 (10%) | >0.9 |
| **Impacted religious practice** | 274 | 23 (8.4%) | 4 (4.3%) | 8 (7.5%) | 11 (15%) | 0.040 |
| *^1^* n (%) | | | | | | |
| *^2^* Pearson’s Chi-squared test; Fisher’s exact test | | | | | | |

**S15 Appendix Table 2: Crude and adjusted estimates for symptom severity score on daily activity impacts.**

Results of logistic regression models investigating the impact of symptom severity scores among contraceptive users in month 3 on whether negative impact on a given symptom was reported in the last month. Adjusted estimates adjust for contraceptive method, urban rural residence, socioeconomic status, parity, occupation, marital status, religion, and education.

| **Activity Type** | **N** | **Crude OR** | **95% CI** | **P value** | **Adjusted OR** | **95% CI** | **P value** |
| --- | --- | --- | --- | --- | --- | --- | --- |
| Job | 116 | 1.00 | 0.93 - 1.06 | 0.950 | **1.23** | **1.07 – 1.45** | **0.007** |
| Education | 60 | 1.02 | 0.89 – 1.23 | 0.781 | 1.13 | 0.80 – 1.65 | 0.494 |
| Chores | **182** | **1.12** | **1.06 - 1.19** | **<0.001** | **1.14** | **1.06 – 1.24** | **0.001** |
| Childcare | **169** | **1.19** | **1.11 – 1.28** | **<0.001** | **1.19** | **1.08-1.35** | **0.001** |
| Relationship harmony | **159** | **1.19** | **1.11 - 1.29** | **<0.001** | **1.18** | **1.07 – 1.33** | **0.002** |
| Religious activities | **180** | **1.22** | **1.13 – 1.32** | **<0.001** | **1.31** | **1.16 – 1.53** | **<0.001** |
