## Supplementary material for "Quantifying Contraceptive Side-Effects: A Prospective Cohort Study of Symptom Burden, Risk Factors, and Daily Life Disruption in South-Central Ethiopia": online supplemental file 15

**S15 Appendix Table: Estimates from minimally-adjusted models for each daily activity investigating the association of each symptom occurrence individually in the third month on whether symptoms were reported to have negatively impacted each daily activity.**

Models adjusted for contraceptive group, residence, socioeconomic status, parity, occupation type, marital status, religion, and education. Symptoms where dashes occur are excluded if there was no one experiencing that symptom among either those who were or were not impacting, mitigating model estimation.

|  | **Job** | | | **Education** | | | **Daily Chores** | | | |
| --- | --- | --- | --- | --- | --- | --- | --- | --- | --- | --- |
| *Predictors* | *Odds Ratios* | *CI* | *p value* | *Odds Ratios* | *CI* | *p value* | *Odds Ratios* | | *CI* | *p value* |
| Increased appetite | 1.27 | 0.33 – 4.89 | 0.726 | 0.11 | 0.01 – 1.94 | 0.131 | 1.67 | | 0.63 - 4.41 | 0.304 |
| Decrease appetite | 1.13 | 0.22 – 5.94 | 0.885 | 0.53 | 0.01 – 36.54 | 0.768 | 2.91 | | 0.80 - 10.51 | 0.104 |
| Fever | 0.58 | 0.10 – 3.50 | 0.549 | 0 | 0 - Inf | 0.995 | 0.82 | | 0.21 - 3.24 | 0.781 |
| Increased blood pressure | - | - | - | - | - | - | 7.90 | | 0.35 - 178.12 | 0.193 |
| Fatigue | 0.79 | 0.27 – 2.35 | 0.668 | 4.97 | 0.37 – 66.40 | 0.226 | 0.74 | | 0.29 - 1.89 | 0.534 |
| Dizziness | **9.34** | **1.77 – 49.43** | **0.009** | 4.7 | 0.23 – 94.60 | 0.315 | **2.65** | | **1.04 - 6.75** | **0.042** |
| Sleep disturbance | 2.79 | 0.60 – 13.02 | 0.191 | >100 | 0 - Inf | 0.997 | **4.88** | | **1.46 - 16.31** | **0.010** |
| Back and joint pain | 1.47 | 0.55 – 3.93 | 0.439 | 1.07 | 0.19 – 5.92 | 0.941 | 1.25 | | 0.53 - 2.94 | 0.604 |
| Arm pain | 0.55 | 0.01 – 3.04 | 0.493 | >100 | 0 - Inf | 0.996 | **8.11** | | **2.61 - 25.18** | **<0.001** |
| Face marks | 0.87 | 0.17 – 4.33 | 0.862 | 0.28 | 0.02 – 3.13 | 0.300 | 1.05 | | 0.19 - 5.91 | 0.959 |
| Gained weight | 0.66 | 0.10 – 4.38 | 0.663 | 1.73 | 0.03 – 99.44 | 0.791 | 2.03 | | 0.54 - 7.63 | 0.296 |
| Lost weight | **5.33** | **1.24 – 22.90** | **0.025** | >100 | 0 - Inf | 0.996 | **4.19** | | **1.35 - 13.02** | **0.013** |
| Not having a period | **3.08** | **1.17 – 8.14** | **0.023** | 0.29 | 0.05 – 1.68 | 0.166 | 1.04 | | 0.43 - 2.52 | 0.923 |
| Nervousness | 1.22 | 0.37 – 4.00 | 0.747 | **0.05** | **0 – 0.636** | **0.021** | 2.26 | | 0.92 - 5.59 | 0.077 |
| Nausea | 2.24 | 0.59 – 8.58 | 0.238 | 0.65 | 0.05 – 9.32 | 0.751 | 0.63 | | 0.18 - 2.20 | 0.473 |
| Headache | 1.14 | 0.40 – 3.21 | 0.806 | >100 | 0 - Inf | 0.995 | 1.14 | | 0.46 - 2.86 | 0.779 |
| Acne | 0.90 | 0.22 – 3.63 | 0.881 | 0.61 | 0.07 – 5.55 | 0.662 | **3.94** | | **1.04 - 14.98** | **0.044** |
| Tender breasts | - |  | - | - | - | - | 10.33 | | 0.74 - 143.48 | 0.082 |
| Abdominal pain | 1.72 | 0.54 – 5.47 | 0.359 | >100 | 0 - Inf | 0.995 | **6.48** | | **2.37 - 17.73** | **<0.001** |
| Abdominal bloating | 0.59 | 0.09 – 3.80 | 0.581 | >100 | 0 - Inf | 0.997 | 1.31 | | 0.32 - 5.34 | 0.705 |
| Discharge | 1.69 | 0.31 – 9.38 | 0.547 | >100 | 0 - Inf | 0.996 | 0.69 | | 0.12 - 3.85 | 0.672 |
| Heavy bleeding | 1.56 | 0.25 – 9.90 | 0.637 | >100 | 0 - Inf | 0.997 | 0.45 | | 0.09 - 2.34 | 0.345 |
| Prolonged bleeding | 0.50 | 0.10 – 2.52 | 0.402 | 13.81 | 0.17 – 1142.53 | 0.244 | 0.49 | | 0.12 - 1.94 | 0.306 |
| Intramenstrual bleeding | 0.30 | 0.03 – 3.17 | 0.320 | >100 | 0 - Inf | 0.997 | 0 | | 0 - Inf | 0.988 |
| Reduced sex drive and enjoyment | 3.20 | 0.74 – 13.82 | 0.120 | 0.15 | 0.01 – 2.61 | 0.196 | **4.59** | | **1.61 - 13.07** | **0.004** |
|  | **Childcare** | | | **Relationship harmony** | | | **Religious activities** | | | |
| *Predictors* | *Odds Ratios* | *CI* | *p value* | *Odds Ratios* | *CI* | *p value* | *Odds Ratios* | *CI* | | *p value* |
| Increased appetite | 1.68 | 0.48 - 5.92 | 0.419 | 1.02 | 0.29 - 3.60 | 0.976 | 0.82 | 0.22 - 3.00 | | 0.759 |
| Decrease appetite | **4.71** | **1.10 - 20.25** | **0.037** | 3.16 | 0.62 - 16.11 | 0.166 | 1.90 | 0.41 - 8.70 | | 0.410 |
| Fever | 0.74 | 0.12 - 4.45 | 0.744 | 1.02 | 0.18 - 5.87 | 0.985 | 0.38 | 0.04 - 3.44 | | 0.392 |
| Increased blood pressure | >100 | 0 - Inf | 0.991 | >100 | 0 - Inf | 0.991 | 0 | 0 - Inf | | 0.993 |
| Fatigue | 0.60 | 0.17 - 2.05 | 0.412 | 2.45 | 0.75 - 8.04 | 0.139 | 1.86 | 0.58 - 5.93 | | 0.297 |
| Dizziness | 2.93 | 0.89 - 9.69 | 0.078 | 2.38 | 0.74 - 7.68 | 0.148 | 3.28 | 0.98 - 10.98 | | 0.055 |
| Sleep disturbance | 3.81 | 0.94 - 15.45 | 0.061 | **10.89** | **2.05 - 57.87** | **0.005** | 4.15 | 0.92 - 18.79 | | 0.065 |
| Back and joint pain | 1.12 | 0.35 - 3.59 | 0.853 | 2.37 | 0.79 - 7.14 | 0.125 | **3.90** | **1.24 - 12.23** | | **0.020** |
| Arm pain | **37.10** | **6.01 - 229.22** | **<0.001** | 3.70 | 0.96 - 14.32 | 0.058 | **7.96** | **1.79 - 35.37** | | **0.006** |
| Face marks | 0 | 0 - Inf | 0.992 | 0 | 0 - Inf | 0.992 | 2.49 | 0.37 - 16.83 | | 0.350 |
| Gained weight | 4.12 | 0.89 - 19.20 | 0.072 | **6.86** | **1.22 - 38.53** | **0.029** | 2.60 | 0.55 - 12.37 | | 0.231 |
| Lost weight | **6.32** | **1.55 - 25.78** | **0.010** | 1.84 | 0.45 - 7.59 | 0.399 | 1.86 | 0.45 - 7.69 | | 0.392 |
| Not having a period | **6.40** | **1.64 - 24.89** | **0.007** | 3.09 | 3.09 - 9.84 | 0.056 | 2.78 | 0.85 - 9.10 | | 0.092 |
| Nervousness | **4.07** | **1.19 - 13.90** | **0.025** | **7.56** | **2.27 - 25.20** | **0.001** | **5.46** | **1.69 - 17.63** | | **0.005** |
| Nausea | 1.01 | 0.24 - 4.27 | 0.994 | 0.61 | 0.13 - 2.93 | 0.541 | 2.62 | 0.73 - 9.40 | | 0.141 |
| Headache | 1.24 | 0.37 - 4.13 | 0.729 | 1.47 | 0.46 - 4.67 | 0.515 | 1.78 | 0.56 - 5.68 | | 0.327 |
| Acne | 0 | 0 - Inf | 0.991 | 0.57 | 0.05 - 5.88 | 0.633 | 2.15 | 0.34 - 13.85 | | 0.420 |
| Tender breasts | 2.74 | 0.15 - 51.58 | 0.501 | **83.28** | **3.57 - >1000** | **0.006** | 6.40 | 0.27 - 152.37 | | 0.251 |
| Abdominal pain | **10.99** | **2.45 - 49.43** | **0.002** | **4.83** | **1.40 - 16.61** | **0.013** | **12.99** | **3.25 - 51.91** | | **<0.001** |
| Abdominal bloating | 1.08 | 0.16 - 7.29 | 0.935 | 2.78 | 0.61 - 12.75 | 0.189 | 4.66 | 0.92 - 23.60 | | 0.063 |
| Discharge | 0.18 | 0.18 - 1.95 | 0.159 | 0.17 | 0.01 - 1.92 | 0.151 | 1.37 | 0.22 - 8.38 | | 0.732 |
| Heavy bleeding | 0.63 | 0.09 - 4.44 | 0.639 | 2.37 | 0.48 - 11.70 | 0.290 | **7.31** | **1.75 - 30.59** | | **0.006** |
| Prolonged bleeding | 0.30 | 0.05 - 1.93 | 0.205 | 2.50 | 0.59 - 10.59 | 0.215 | 3.28 | 0.91 - 11.79 | | 0.069 |
| Intramenstrual bleeding | 0 | 0 - Inf | 0.992 | 0.28 | 0.02 - 3.55 | 0.323 | 0.65 | 0.09 - 4.69 | | 0.668 |
| Reduced sex drive and enjoyment | **10.66** | **2.54 - 44.68** | **0.001** | **73.98** | **11.27 - 485.53** | **<0.001** | **18.77** | **4.51 - 78.11** | | **<0.001** |
